# All-cause, circulatory disease and cancer mortality in the population of a large Italian area contaminated by perfluoroalkyl and polyfluoroalkyl substances (1980-2018)

**DOI:** 10.1101/2023.12.09.23299764

**Authors:** Annibale Biggeri, Giorgia Stoppa, Laura Facciolo, Giuliano Fin, Silvia Mancini, Valerio Manno, Giada Minelli, Federica Zamagni, Michela Zamboni, Dolores Catelan, Lauro Bucchi

## Abstract

**Background:** Perfluoroalkyl substances (PFAS) are associated with many adverse health conditions. Among the main effects is carcinogenicity in humans, which deserves to be further clarified. A clear association has been reported for kidney cancer and testicular cancer. In 2013, a large episode of surface, ground and drinking water contamination with PFAS was uncovered in three provinces of the Veneto Region (north-eastern Italy) involving 30 municipalities and a population of about 150,000. We report on the temporal evolution of all-cause mortality and selected cause-specific mortality by calendar period and birth cohort in the local population between 1980 and 2018.

**Methods:** The Italian National Institute of Health and the Italian National Institute of Statistics made available the anonymous records of death certificates of residents of the provinces of Vicenza, Verona and Padova (males, n=29,629; females, n=29,518) who died between 1980 and 2018. Calendar period analysis was done by calculating standardized (European standard population 2013) mortality ratios using the total population of the three provinces in the same calendar period as a reference. The birth cohort analysis was performed using cumulative risks for ages 20-84 years.

**Results:** During the 33 years between 1985 (assumed as beginning date of water contamination) and 2018 (last year of available cause-specific mortality), in the resident population of the *Red area* (the municipalities being connect to the contaminated water plant) we observed 51,621 deaths as compared to 47,731 expected (age-sex SMR: 108; 90% CI: 107-109). We found evidence of raised mortality for diseases of the circulatory system (in particular, heart disease and ischemic heart disease) and malignant neoplastic diseases, including kidney cancer and testicular cancer.

**Conclusions:** For the first time, an association of PFAS exposure with mortality from circulatory disease mortality was formally demonstrated. Circumstantial evidence regarding kidney cancer and testicular cancer is consistent with previously reported data.

## Background

The synthetically-made industrial chemicals per-and polyfluoroalkyl substances (PFAS) have been marketed since the 1940s and have been extensively used since the 1950s in a wide variety of applications [1]. According to the U.S. National Institute of Environmental Health Sciences, there are some 15,000 different PFAS molecules, but hundreds of newer and previously unreported PFAS and PFAS-related compounds –the so called “emerging” PFAS– continue to be identified by researchers [2, 3].

PFAS have the unique property of a hydrophobic-lipophilic carbon chain with fluorine atoms rather than hydrogen atoms in at least part of the carbon atoms. Since the carbon-fluorine bond is among the strongest ones, PFAS have a high resistance to degradation. This makes them to be industrially valuable, especially in applications at high temperatures or high pressures or in corrosive environments [1–3].

Closely linked to this industrial success, however, there is the problem that PFAS are difficult to break down through normal chemical, physical or biological processes, including conventional water/wastewater treatment processes [4]. This causes their persistence in the soil, water, air and organisms [5]. PFAS have been found all over the world in a variety of environmental media [4], including surface, ground and drinking water [6–8]. Quantitatively, manufacturing facilities are the most important sources of contamination [9, 10] followed by emissions from municipal and industrial wastewater treatment plants [9, 11–13].

People are most commonly exposed to PFAS by consuming contaminated water or food (including contamination from food packing), or using products made with PFAS, or breathing PFAS-contaminated air. Given their persistence, blood levels of some PFAS may increase over long periods of time [14]. Increasing epidemiologic research has suggested that exposure to high levels of PFAS might be associated with many adverse health conditions, including decreased fertility in men [15] and women [14], birth defects [16], delayed development [17], osteoporosis in young men [18], damage to the immune system [19], reduced antibody response after vaccination, allergy and asthma in children [20], liver disease [21], increased levels of serum cholesterol [22], impaired thyroid function [23], insulin resistance [24], gestational diabetes [25], pregnancy-induced hypertension [26], neuro-endocrine disruption [27] and cancer.

With respect to the latter, the International Agency for Research on Cancer has re-evaluated the carcinogenicity of PFOA and PFOS in November 2023 [28]. PFOA has been classified as ‘carcinogenic to humans’ (Group 1) based on ‘sufficient’ evidence in experimental animals and ‘strong’ mechanistic evidence in exposed humans indicating an association with epigenetic alterations and an immunosuppressive action. In addition, there is ‘limited’ epidemiologic evidence in humans for an association with kidney cancer and testicular cancer. PFOS has been classified as ‘possibly carcinogenic to humans’ (Group 2B) based on ‘strong’ mechanistic evidence. The evidence for cancer in experimental animals has been considered to be ‘limited’ and the evidence regarding cancer in humans ‘inadequate’. This is due to the fact that the few available studies have led to positive findings only sporadically and inconsistently. It appears that there remains much room for further epidemiologic research.

An ever-increasing number of contaminated sites are being discovered in many countries [29]. The most important evidence regarding PFAS exposure has come from a series of studies investigating the community living near the DuPont Washington Works fluoroproduct manufacturing facility in Parkersburg, West Virginia, U.S. [30–33]. However, the world’s largest episode of PFAS water-contamination reported so far occurred in Italy [34]. In 2011, the Institute of Water Research of the National Research Council (IRSA– CNR) was commissioned a study by the Italian Ministry for the Environment to investigate the contamination with PFAS in the most important river basins of the Country [35]. In 2013, the study revealed the surface, ground and drinking water contamination of an area of the Veneto Region (northeastern Italy) involving a total of about 125.000 persons according to early estimates [13, 34].

In the present article, we report a study on the temporal evolution of all-cause mortality and selected cause-specific mortality by calendar period and birth cohort in the population living in the contaminated area between 1980 and 2018.

## Methods

### Study setting: environmental monitoring and source, area and timing of contamination

#### Environmental monitoring plan

The results of the study on the contamination with PFAS in the Italian river basins [36] were communicated to the Veneto Regional Administration in the summer of 2013 [37]. Soon after, the Environmental Prevention and Protection Agency of the Veneto Region (ARPAV) established an environmental monitoring plan, which is still operating, with the two-fold objectives of (1) assessing the geographic extension and the level of groundwater and drinking water contamination with PFAS and (2) identifying its source(s). Originally, 12 types of PFAS were being measured in water matrices, namely: perfluorobutanoic acid (PFBA), perfluorobutanesulfonic acid (PFBS), perfluoropentanoic acid (PFPeA), perfluorohexanoic acid (PFHxA), PFHxS, perfluoroheptanoic acid (PFHpA), PFOA, perfluorooctane sulfonic acid (PFOS), perfluorononanoic acid (PFNA), perfluorodecanoic acid (PFDeA), perfluoroundecanoic acid (PFUnA), and perfluorododecanoic acid (PFDoA) [34].

Chemical analysis of drinking water samples during 2013 indicated the presence of PFOA (median, 319 ng/L), PFBA (123 ng/L), PFBS (91 ng/L), PFPeA (70 ng/L), (PFHxA (52 ng/L), PFOS (18 ng/L), PFHpA (14 ng/L) and PFHxS (<10 ng/L) [34]. Further details can be found in another publication [38].

#### Source of contamination with PFAS

The MITENI (formerly RIMAR) factory, a manufacturing plant located in the town of Trissino (province of Vicenza), has been identified as the most likely source of contamination with PFAS in the region. The factory was established in the 1950s. The production of PFAS was started in 1966 and was terminated in 2018 [39]. Long-chain PFAS (containing ≥ 6 carbons, such as the PFOA, or ≥ 7 carbons, such as the perfluoroalkyl carboxylic acids) were produced between 1968 and 2001, short-chain PFAS (for example, PFBA) between 2013 and 2016, and a mixture of molecules between 2001 and 2013 [39].

The MITENI factory discharged the treated effluent into the municipal wastewater treatment plant, whose output is mixed with the outputs of other four plants and carried by a single sewer pipeline (known as *collettore ARICA*) to the Fratta-Gorzone river. These five plants collect waters from domestic sewage and industrial districts of the province of Vicenza. This makes it impossible to distinguish the effluent originating from the fluorochemical plant from those discharged by the other industrial activities (mainly textile and tannery factories). However, a comparison between the PFAS composition of the primary discharge of the fluorochemical plant and that of the output of the *collettore ARICA* revealed a close similarity [13, 36].

#### Area of water contamination with PFAS

The contamination of surface and deep waters took place in the area located downstream of the MITENI factory, which has a well-developed agricultural and food industry. The groundwater contamination plume occupies a surface area of 190 square kilometers and involves public aqueducts and private wells across the provinces of Vicenza, Verona, and Padova. Based on the results of the environmental monitoring, the Environmental Protection Agency of Veneto identified 30 municipalities forming an area of maximum exposure (*Red area*), for a population of 153,525 inhabitants in 1 January 2020 [40], a larger number than previously estimated [13, 34]. The *Red area* is further divided into *Red area A*, including the municipalities that are served by the contaminated aqueduct and are also located on the groundwater contamination plume, and *Red area B*, including municipalities served by the contaminated aqueduct but not located on the groundwater contamination plume.

The contamination has been mapped by the Veneto Regional Administration in 2013. The maps are available online [41]. A digital navigable map is available from another website [42]. Fig. 1 shows the map of water contamination by PFOA and PFOS in the years 2013-2015, i.e. when the contamination was discovered and before the full implementation of mitigating or remediation activities. In the map, the 30 municipalities of the *Red Area* served by the aqueduct fed by the contaminated groundwater since 1984 are also shown.

**Figure 1.**
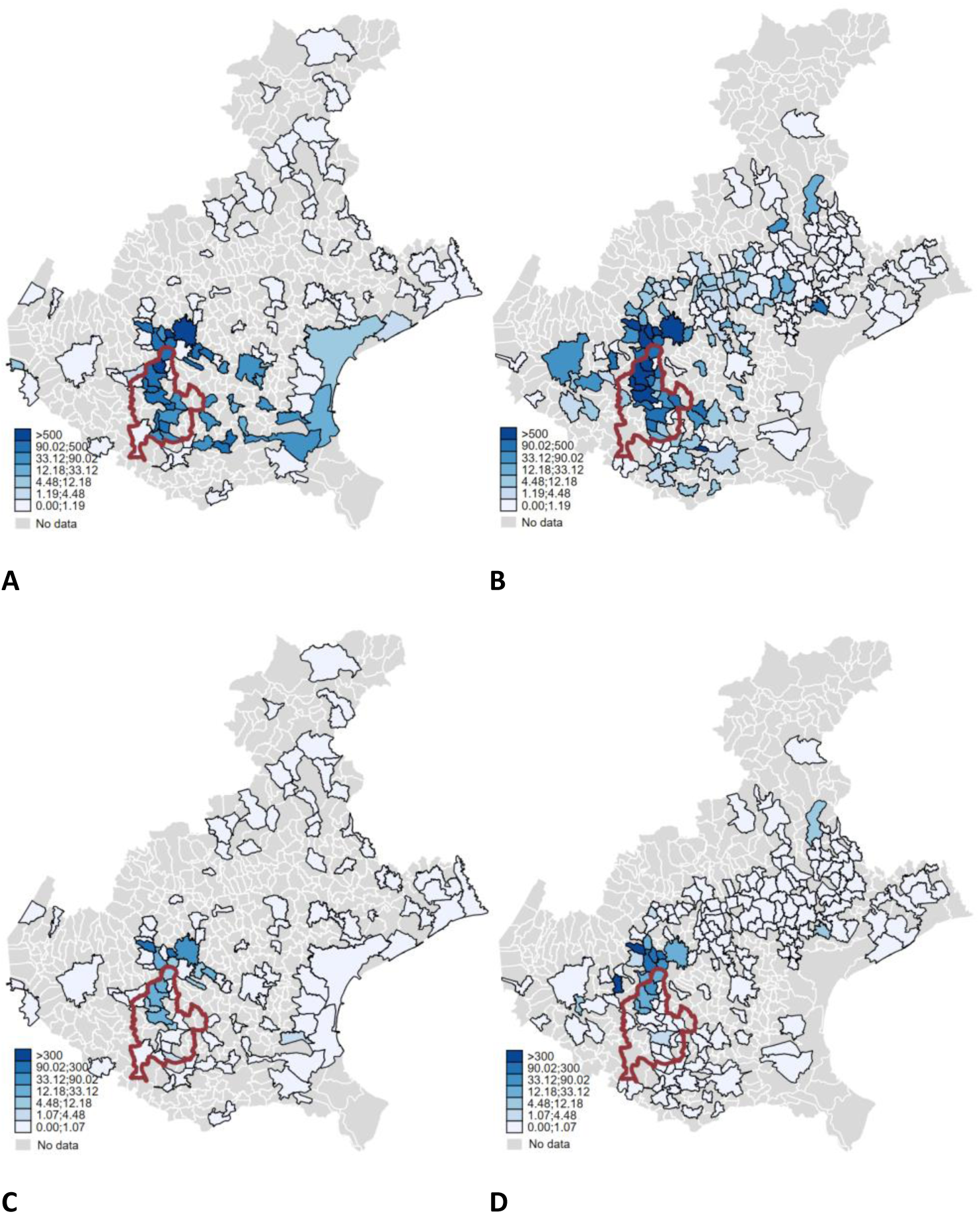
Map of the concentrations (ng/l) of perfluorooctanoic acid (PFOA) (top panel A and B) and perfluorosulfonic acid (PFOS) (bottom panel C and D) in the shallow water (left panel A and C) and the groundwater (right panel B and D) of the Veneto Region municipalities. July 2013–June 2015. Source Environmental Protection Agency Veneto Region (ARPAV) https://www.arpa.veneto.it/dati-ambientali/open-data/idrosfera/concentrazione-di-sostanze-perfluoroalchiliche-pfas-nelle-acque-prelevate-da-arpav accessed october 18th, 2023

In this study, we defined the exposed population as the resident population of the *Red Area. The Red area A* includes the municipalities of (in parentheses, the province to which the municipality belongs) Alonte (Vicenza, VI), Asigliano Veneto (VI), Brendola (VI), Lonigo (VI), Sarego (VI), Noventa Vicentina (VI), Orgiano (VI), Pojana Maggiore (VI), Montagnana (Padova, PD), Cologna Veneta (Verona, VR), Pressana (VR), Roveredo di Guà (VR), and Zimella (VR). The municipalities situated in the *Red area B* are Albaredo d’Adige (VR), Arcole (VR), Bevilacqua (VR), Bonavigo (VR), Boschi Sant’Anna (VR), Legnago (VR), Minerbe (VR), Terrazzo (VR), Veronella (VR), and Urbana (PD). The *Red area B* also includes parts of the municipalities of Agugliaro (VI), Borgo Veneto (PD), Casale di Scodosia (PD), Lozzo Atestino (PD), Megliadino San Vitale (PD), Merlara (PD), and Val Liona (VI).

#### Timing of water contamination with PFAS

According to estimates from the ARPAV, the contamination of the groundwater began in 1966 in Trissino [43] and reached Montecchio Maggiore in 1970, Almisano in 1984 and Lonigo –situated further south– in 1985.

In the period 1985-1989, unfortunately, several aqueducts drew water the Almisano groundwater source and, in the same area, a large aqueduct was built (*Centrale Madonna di Lonigo*, completed in 1995). This delivers drinking water to the 30 municipalities in the *Red area*. Thus, the whole *Red area* was contaminated since the years 1985-1989 [43].

PFAS exposure via drinking water ingestion from the public aquifer serving the *Red area* has been reduced since 2013, with the implementation of a filtration system with granular activated carbon. The levels of specific PFAS were found to be below the level of detectability of 5 ng/L in 2018. The population served by private wells for drinking water, which lack filtration systems, was estimated at around 18,000 people in 2016.

#### Food contamination with PFAS

In 2016, the Veneto Regional Administration developed a plan for the monitoring of 12 PFAS congeners in local food matrices. After the implementation, the Italian National Health Institute (ISS) performed a scenario-based estimate of the weekly dietary exposure in the *Red area* [44]. In 2021, food contamination and food contribution to total human exposure were assessed using the individual records of the food monitoring campaigns of the years 2016-2017. The analysis showed that food PFAS contamination was widespread over the entire *Red area* with modest differences [45]. The exportation of local food may have caused exposures outside the *Red area*.

#### Health surveillance plan

Between July 2015 and April 2016, the ISS performed the first serum PFAS concentration measurements campaign in the local population [46]. PFOA had the highest concentration but detectable levels of eight other molecules (PFBA, PFPeA, PFHxA, PFHpA, PFDoA, PFBS, PFHxS, and PFOS) were found. The median serum level of PFOA was 14 (95th percentile: 248 ng/mL). For PFBA, the median concentration was below detection and the 95th percentile was 0.6 ng/mL), reflecting its more rapid excretion.

In 2017, the Veneto Regional Administration developed a health surveillance plan for the exposed population [34, 47]. The implementation started in 2017-2018 from the birth cohorts born in 1962-2002 and was subsequently extended to include the other birth cohorts. The plan is based on active personal invitations to undergo measurements of PFAS in serum, laboratory tests and medical examinations mainly to assess serum lipids as well as liver and kidney functionality, followed by specialist diagnostic work-up, if necessary. The plan covers the 30 municipalities of the *Red area*.

In Supplementary Tables S1-S2, the mean, the median and the 95th percentile of serum levels of PFOA and PFOS by attained age at first invitation (range age class: 7-72 years) and year of invitation (2017-2023) to the health surveillance plan are shown [47].

### Demographics, migration balance and deprivation index

The background demographic figures of the three reference provinces showed a smooth increase between 1981 and 1991, a steeper growth since 1991 and a plateauing since 2011 (Supplementary Figure S1). There was no appreciable difference between the three provinces and the municipalities of the *Red area*. Peculiar patterns were observed only in the municipalities of Lonigo, where a marked increase occurred between 1981 and 2011, and Montagnana, which showed a decreasing trend since 1981.

The natural balance and the net migration in the years 2003-2017 were consistent with the above patterns, with Lonigo showing a positive balance and Montagnana a negative balance. As expected, the three provincial capital cities of Vicenza, Verona and Padova exhibited a strong negative trend of the natural balance, with a positive net migration occurring in Verona and Padova. (Supplementary Figures S2-S3)

As shown in the age-sex pyramids (Supplementary Figure S4), the age distribution in 1982 and 2018 did not differ between the three provinces and the 30 municipalities of the *Red area*. In the latter, it appears that the maximum impact of the PFAS contamination is experienced by the cohorts born between 1960 and 1970, who were aged 10-20 years in 1982.

The socioeconomic characteristics of the municipalities of the *Red area* did not differ much from the reference populations of the three provinces. In 2011, the material deprivation index was similar (Supplementary Figure S5). The analysis of five major socio-economic characteristics pertaining to deprivation indicators (low education (<6yrs), unemployment, house crowding, house tenure, and single parent families) showed only a relatively larger percentage of low-educated people (32% versus 30%) in the *Red area* population (Table 1). A more detailed picture can be found in the biplot reported in Supplementary Figure S6.

**Table 1.**
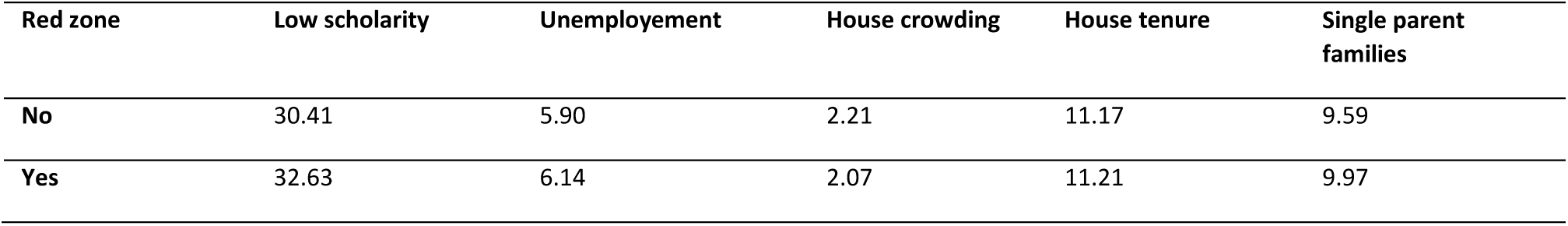
Percentage of people with low scholarity, unemployment, house crowding, house tenure and single parent families in the three provinces of Vicenza, Verona and Padova and the 30 Municipalities of the PFAS contaminated Red Area.

| Red zone | Low scholary | Unemployment | House crowding | House tenure | Single parent families |
| --- | --- | --- | --- | --- | --- |
| No | 30.41 | 5.90 | 2.21 | 11.17 | 9.59 |
| Yes | 32.63 | 6.14 | 2.07 | 11.21 | 9.97 |

### Study rationale and design

The rationale of this study is based on the consideration that the PFAS contamination that occurred in the Veneto Region is extremely important in terms of size of the exposed population and magnitude of exposure. The accident can be taken as a great and tragic *natural* experiment [48]. PFAS exposure of the population began in the second half of the 1980s and depended on the use of contaminated water. The experimental condition consists in being connected to the aqueduct, which acted as an instrumental variable. The actual PFAS exposure could depend on several factors and, among these, on the determinants of contaminated water consumptions. The relationship between being connected to the aqueduct and a disease outcome was not affected by confounding, while the relationship between actual PFAS exposure and a disease outcome could be confounded. An instrumental variable should be unrelated to the outcome, strongly related to the exposure and unrelated to the potential confounders. Being connected to the aqueduct was strongly associated to the exposure and was independent from potential confounders and –of course– from the outcome. As a consequence, comparing the population connected to the aqueduct with an unconnected reference population enables to test a potential causal association. Of course, we should consider the possibility of residual confounding in the definition of the two populations and of potential misclassification, because we had only information at the municipality level. On average, more than 70% of the population of the *Red Area* was served by the aqueduct. In *Red Area A,* furthermore, the population not served by the aqueduct was supplied by private wells containing the contaminated groundwater.

The potential fallacy connected to the use of the information on PFAS exposure at the population level for the definition of the groups being compared can be exemplified by considering the municipality of Lonigo. This municipality has shown a higher serum PFAS concentration in the health surveillance plan and has some specific socioeconomic characteristics that are strongly linked to the volume of water consumption [49, 50] and cardiovascular mortality [51] like education and house tenure. This causes socio-economic factors to act as confounders of the association between PFAS exposure and cardiovascular disease (Supplementary Figure S7).

The availability of mortality data for a time span as long as 1980-2018 enabled a temporal analysis of mortality risk by two different time axes. The temporal variations of mortality were evaluated both by focusing on the effect of calendar period and by investigating the birth cohort influences. The effect of calendar period is interpretable as the effect of the total dose of PFAS since 1984. Therefore, mortality in the 1980-1984 calendar period can be assumed as an internal reference, with the subsequent calendar periods being characterized by a mortality experience that reflects growing exposure doses. The effect of birth cohort, conversely, is interpretable as the effect of the exposure which begins at different ages. Therefore, the exposure started after at least 60 years of age for the cohort born in 1920-1929 and after 25 years of age for the cohort born in 1960-1969.

The bioaccumulative nature of PFAS led to a constantly increasing exposure of the population. Since water filtration was implemented starting from 2013, it could potentially impact the last time segment, i.e. 2015-2018. These public health measures, coupled with the health surveillance activities and changes in lifestyle and health behaviour after the discovery of water contamination, may have contributed to a reduction of PFAS exposure of the population.

We used the population of the three provinces where the *Red area* is situated as a reference population. In a sensitivity analysis, we used as reference the population of the tree provinces excluding the 30 municipalities of the *Red area* (the results –not shown– are available from the corresponding author upon request).

### Source of mortality data

The Italian National Institute of Health (ISS) made available to us the anonymous records of death certificates of all males (n = 29629) and females (n = 29518) who were residents of the provinces of Vicenza, Verona and Padova and died between 1980 and 2018 (more recent data were not yet ready for use at the time of analysis). Records were extracted by the Statistical Service of the ISS from The Italian National Institute of Statistics cause-specific mortality database using the codes from the International Classification of Diseases (ICD)-9 [52] between 1995 and 2002 and ICD-10 [53] thereafter. The ICD-9 codes have been converted into ICD-10 codes. (Supplementary Table S3).

In order to facilitate other comparisons, observed deaths, directly standardized mortality rates (2013 European standard population) per 100,000, 0-59 and 60-85 cumulative mortality rates, attributable deaths and probability of 0 attributable deaths are provided in Supplementary Table S4.

### Statistical analysis

Data were aggregated into 18 five-year age groups (0-4, 5-9, …, ≥85) and 8 five-year periods (with the exception of the last one which is a four-year period: 1980-1984, 1985-1989, 1990-1994, 1995-1999, 2000-2004, 2005-2009, 2010-2014,…, 2015-2018). Figure 2 shows the Lexis diagram and the reconstruction of birth cohorts. The 15 cohorts that were considered in the analysis are highlighted.

**Figure 2.**
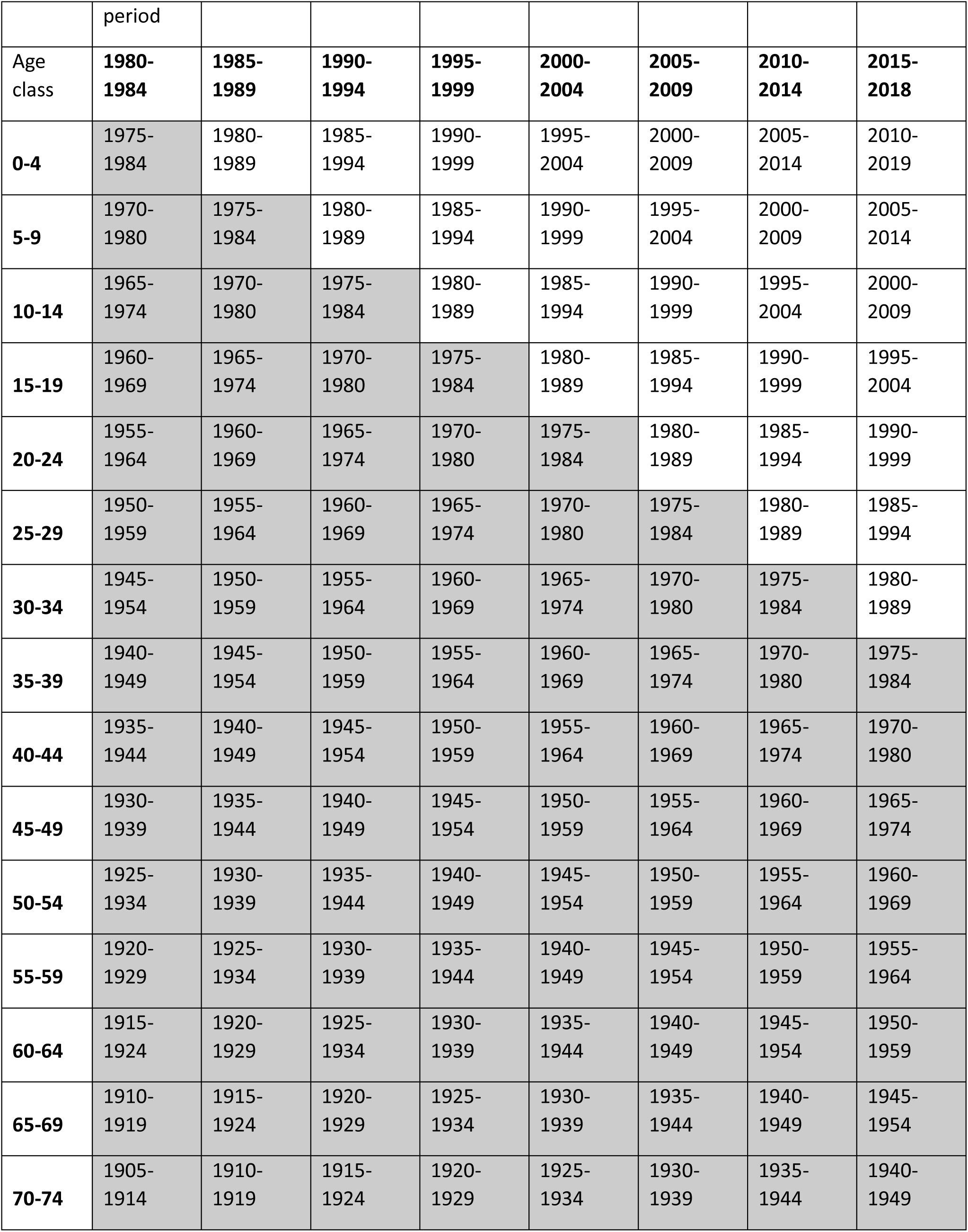
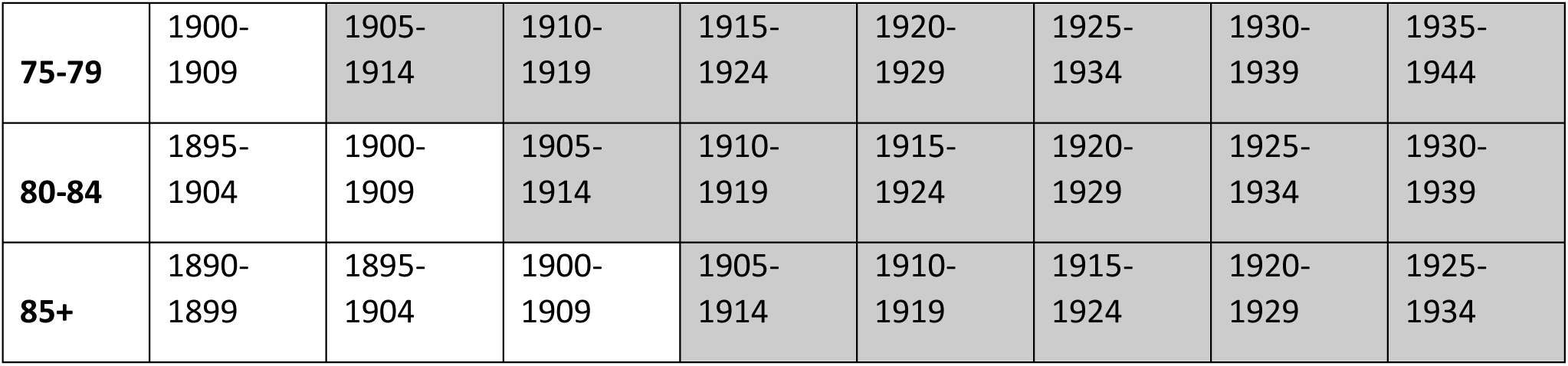
Lexis diagram showing age classes, calendar period and highlighted in gray the analyzed birth cohorts

Calendar period analysis was carried out calculating direct standardized rates (European standard population 2013) and the standardized mortality ratios (SMRs) using the rates observed in the populations of the three provinces to which the 30 municipalities belong (Vicenza, Verona and Padova) for the same calendar period as reference.

The birth cohort analysis were performed using cumulative risks 20-84 (CR) years and cumulative SMRs (SMRcum) 20-84 years.

Let define O_𝑖c_ and 𝑃*Y*_ic_ the number of observed deaths and person years of observation for the i-th age class and c-th birth cohort. We fitted a log-linear Poisson regression model:

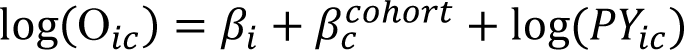

where 𝛽_𝑖_ is the coefficient for the i-th age-class (logarithm of the specific i-th age-class rate), 𝛽*_c_^cohort^* the coefficient for the c-th cohort (logarithm of the ratio between the rate for the c-th cohort and the rate of the reference cohort) and log(𝑃_𝑖_) is the offset, that is, the weight to be attributed to the observed frequency. For the analysis of the age-cohort model, the central cohort or the one with the largest number of complete data is usually used as a reference category and in our case the cohort of those born in the years 1935-1944 was chosen [54, 55]. The intercept is not included in the model, so the age-specific coefficients can all be interpreted directly as logarithms of the specific rates. The cumulative risk 20-84 for the c-th cohort is obtained as

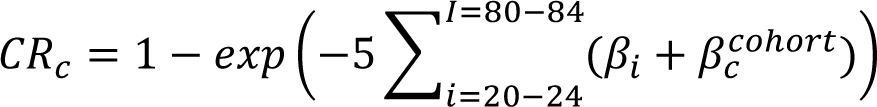

and its confidence interval is obtained through the delta method [56].

We calculated also cumulative the SMR20-84 for the analyzed birth cohorts.

The regression model is similar to the one described above, but with the logarithm of the number of expected cases as offset. For the c-th cohort the cumulative SMR20-84 is given by:

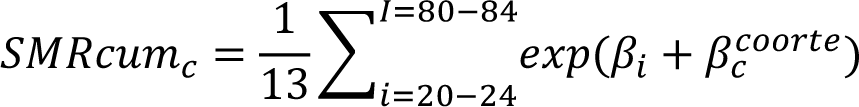

In this case, the number of expected cases was obtained for each calendar period on the basis of the age-specific rates of the reference population [57].

More details can be found in Supplementary Table S4.

The analysis of the trend in mortality from testicular cancer is difficult due to the strong increase in survival since 1980. For Italy, an age-period-cohort analysis showed a striking decrease in mortality after year 2000 [58]. We therefore analyzed mortality for testicular cancer only for the period 1980-1999.

## Results

Overall, during the 33 years from 1985 (assumed date of water contamination) to 2018 (last year of availability of cause-specific mortality), in the resident population of the *Red area* we observed 51,621 deaths over 47,731 expected deaths (according to the mortality rates of the three provinces which include the *Red area* population), i.e. an excess of 3890 deaths (90% CI: 3543-4231). In other words, every three days there were 12 deaths over 11 expected. The overall SMR was 108 (90% CI: 107-109). In Supplementary Table S5, the SMRs 1985-2018 for all the analyzed causes of death are shown. We found evidence for increased mortality from diseases of the circulatory system (particularly heart diseases and ischemic heart diseases), diabetes, diseases of the digestive system, malignant neoplastic diseases (in men), liver cancer (in men), lung cancer (in men), female breast cancer, and kidney cancer. In general, all excesses were larger for the population living in the *Red area A*.

To summarize the time pattern, we compared the SMR for the period 1980-1989 with the overall SMR for the period 1980-2018. This is equivalent to saying that, adopting a conservative approach, we considered the 1980s as a period without considerable population exposure. We also compared the SMR for the last observed period, 2010-2018, with the overall SMR for the years 1980-2018. In other words, we considered the years 2010-2018 as the period with the highest level of cumulative exposure. (Supplementary Table S6).

For all causes of death, we found strong evidence for an increased mortality risk since the baseline level of the 1980s, an increase that was present even after 2010. This pattern was observed both in the *Red Area* and particularly in the *Red Area A* and both in males and females.

Mortality from diseases of the circulatory system, heart diseases and ischemic heart disease showed a comparable trend.

Diseases of the digestive system and diabetes showed a mortality increase after 2010, although this was not observed in the *Red Area A*.

Mortality from malignant neoplasm increased after the 1980s in both sexes. The pattern for specific cancer sites is less clear due to larger statistical uncertainty. We did not find evidence for increasing mortality from liver, lung, breast, ovarian and thyroid cancers and malignant neoplasms of the lymphohematopoietic system. We found evidence for an increased risk of dying from kidney cancer in both sexes and testicular cancer.

The excess risk could also vary by birth cohort, being more severe the younger the age at first exposure. If the time trend is linear, however, we cannot disentangle the effect of calendar period from the effect of birth cohort. Due to statistical consideration regarding the sample size and the number of events, we performed an age-period-cohort analysis on all causes mortality. The results showed a greater importance of the birth cohort time axis among men, while for women we found more relevant the calendar period time axis (see Table 2). In the following we describe the main results in term of birth cohort and then of calendar period.

**Table 2.** Summaries of fitting different age-period-cohort models to all-cause mortality data. Marginals likelihood ratio (LR), degrees of freedom (df). PFAS contaminated Red Area. 1980-2018.

|  | Men | Women |
| --- | --- | --- |
|  | All causes | All causes |
|  | LR (df) | LR (df) |
| Age | 147 (125) | 131 (125) |
| + drift | 144 (124) | 126 (124) |
| drift + cohort | 94 (101) | 105 (101) |
| drift + period | 117 (118) | 109 (118) |
| drift + cohort + period | 87 (95) | 95 (95) |

### Birth cohort analysis

#### All-cause mortality

Figure 3 shows the cumulative SMR for all-cause mortality. Among women, panel B shows that the cumulative SMR for all-cause mortality grew from the cohort born in 1920-1929 and peaked (RR: 1.15; 90% confidence interval (CI): 1.05-1.25) in the birth cohort of 1945-1954, i.e. those women exposed from the age of 35-39 and older. Notably, the SMRs among women aged less than 35 years in 1984-1989, born after 1960, did not differ from the reference population.

**Figure 3.**
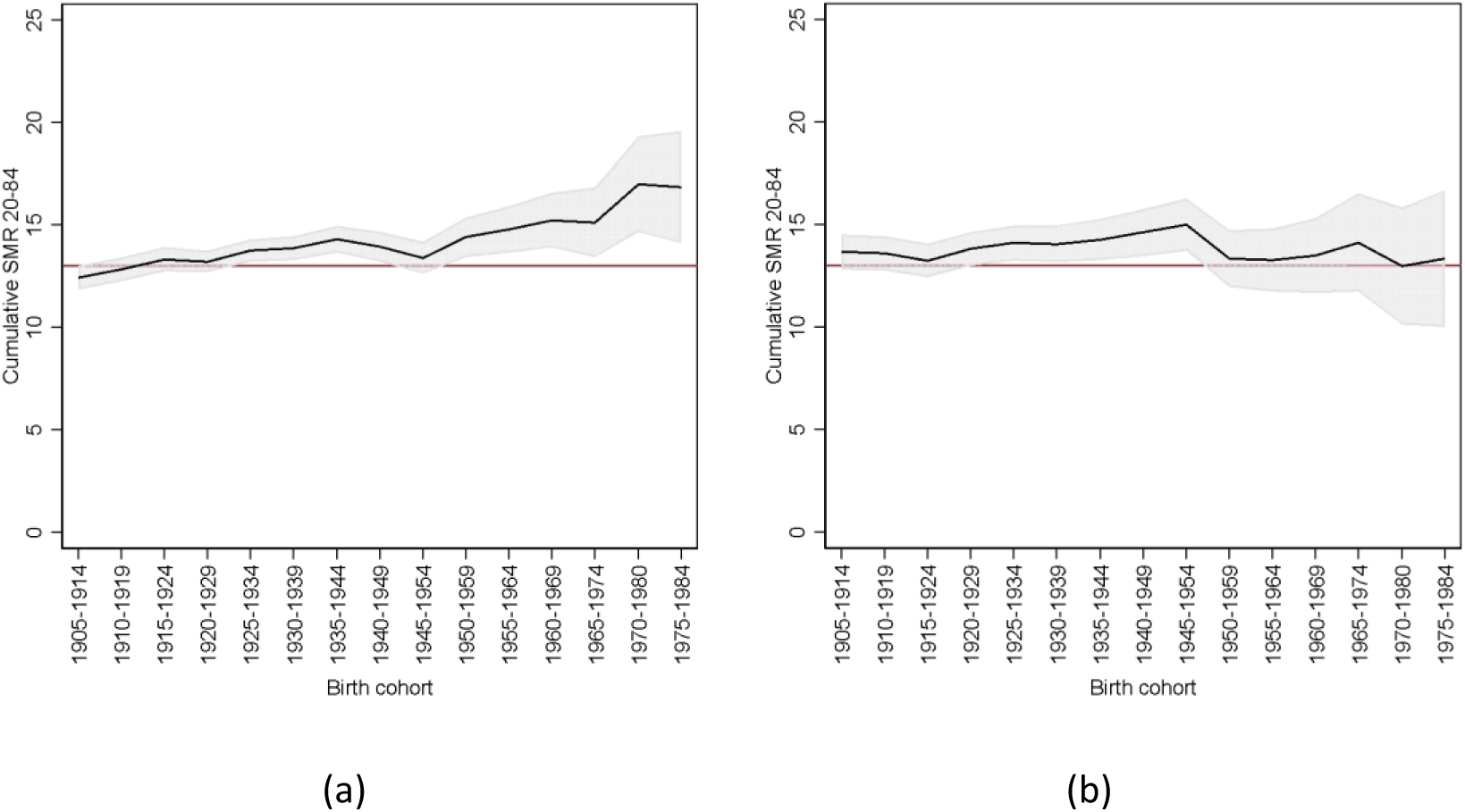
Cumulative SMR (and 90% confidence band, grey area) for all-cause mortality by birth cohort. Males (a) and females (b). PFAS contaminated Red Area. Birth cohorts 1905-1984. (Reference value: red line)

Among men (panel A), the cumulative SMRs rose consistently and almost linearly starting with those born in 1920-1929 and after, with a peak (RR: 1.31; 90% CI: 1.13-1.49) in the birth cohort of 1970-1980. This reflected the growing cumulative exposure to a persistent organic pollutant. The birth cohort dimension, thus, was more important than the calendar period. By the way, comparing the two sexes, the cumulative SMRs among women born until 1945-54 rose more steeply than among men. Among men, the RR of 1.14 (90% CI: 1.05-1.22) was reached by the birth cohort of 1955-1964, i.e. ten years later than women.

#### Circulatory mortality

These patterns are similar for diseases of the circulatory system but with greater statistical uncertainty.

#### Cancer mortality

Figure 4 shows that, among females, the cumulative SMR for mortality from malignant tumours grew starting with the birth cohort of 1945-1954 and peaked (RR: 1.52; 90% CI: 0.85-2.19) in the cohort of those born in the years 1975-1984, i.e. the women exposed from the age of 5-9 years. Among males, the cumulative SMRs rose almost linearly starting with those born in 1935-1944 and after, peaking (RR: 1.35 90% CI: 0.71-1.98) in the birth cohort of 1975-1984, i.e. the men exposed from the age of 5-9 years. Patterns similar to this were also observed for specific tumour sites but with strong statistical uncertainty.

**Figure 4.**
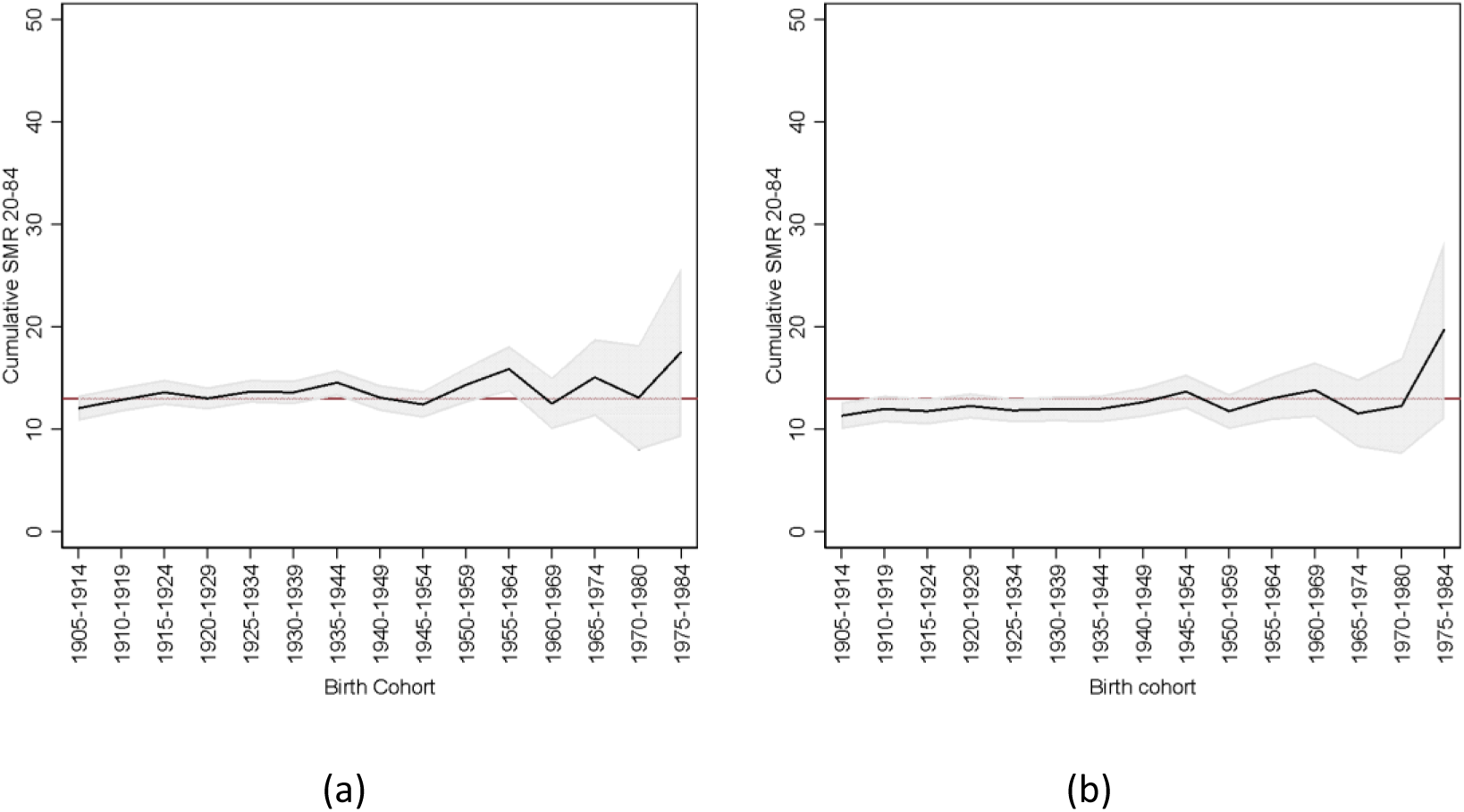
Cumulative SMR (and 90% confidence band, grey area) for malignant tumours mortality by birth cohort. Males (a) and females (b). PFAS contaminated Red Area. Birth cohorts 1905-1984. (Reference value: red line)

### Calendar period analysis

Supplementary Table S5. shows the observed deaths, the SMRs and the 90% CI for each 5-year calendar period between 1980 and 2018, by sex and cause of death. Here we point out the most relevant findings.

#### Circulatory mortality

Figure 5 shows the sex-and cause-specific SMR according to calendar period. The SMR exceeded the null value from the calendar period 1985-1989 onward, in both women and men. There was no evidence for a decrease after 2014. In the calendar period 1980-1984, no excess risk was evident. The SMRs in the population of the *Red area A* were higher, peaking in 2015-2018 at 131 (90% CI: 123-139) for females and 120 (90% CI: 111-129) for males.

**Figure 5:**
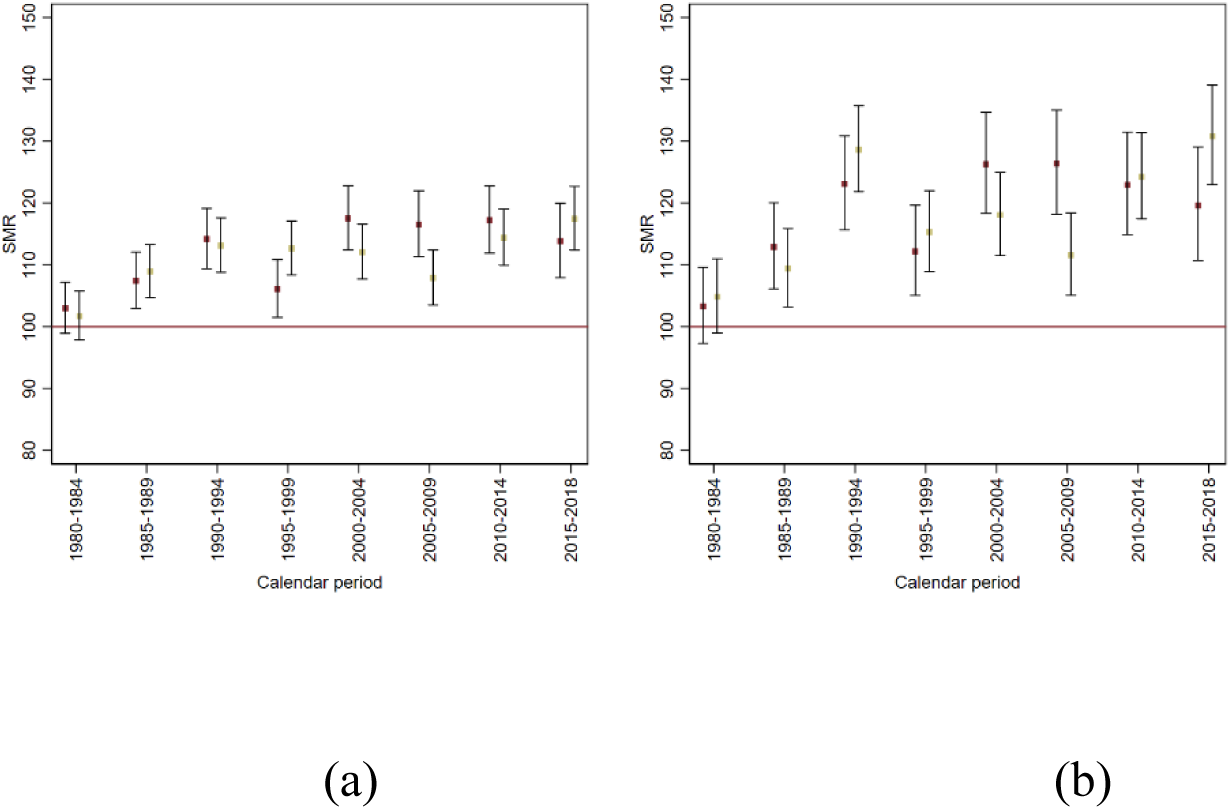
Mortality for diseases of the circulatory system. Sex-specific SMR by calendar period (Khaki Females Red Males). Panel (a) *Red area*; (b) *Red area A*. Red Area refers to the 30 Municipalities of Veneto Region connected to the PFAS contaminated water plant (aquifer), Red Area A refers to the 13 municipalities with also PFAS contamination of groundwater. 1980-2018.

#### Cancer mortality

With respect to malignant neoplasms (Table 3) we found an increasing trend by calendar period in both sexes. Taking the 1980s (1980-1989) as a reference and considering two broad time periods (1990-2009 and 2010-2018), we observed a clear period effect (LR: 10.66; 2 df; p=0.005), without evidence for a difference by sex (LR: 2.18; 2 df; p=0.337). Consistently with the long latency period for cancer, an excess risk was observed more than 20 years after the water contamination (see Table 4 for details).

**Table 3.** Observed and expected deaths of malignant tumours by sex and calendar periods. PFAS contaminated Red Area population. 1980-2018.

| Trend | Males |  | Females |  |
| --- | --- | --- | --- | --- |
|  | Observed | Expected | Observed | Expected |
| 1980-1989 | 2213 | 2243 | 1371 | 1452 |
| 1990-2009 | 4866 | 4866 | 3218 | 3375 |
| 2010-2018 | 2151 | 1981 | 1632 | 1656 |

**Table 4.** Summaries of fitting a poisson regression model on malignant tumours. PFAS contaminated Red Area population. 1980-2018**. IRR: mortality rate-ratio.**

|  | IRR | pvalue | 90% IC |
| --- | --- | --- | --- |
| Female vs. Male | 0.92 | <0.001 | 0.89-0.94 |
| 1980-89 | reference |  |  |
| 1990-2009 vs 1980-1989 | 1.05 | 0.021 | 1.01-1.08 |
| 2010-2018 vs. 1980-1989 | 1.08 | 0.001 | 1.04-1.12 |

#### Kidney cancer mortality

Figure 6 shows the sex and cause-specific SMR according to calendar period. The SMR by calendar period exceeded the null value from 2015-2018 in both women and men. In the period 1980-1984, no excess was evident. Similar considerations apply to kidney cancer mortality figures for the *Red area* overall, as summarized in Table 5.

**Figure 6:**
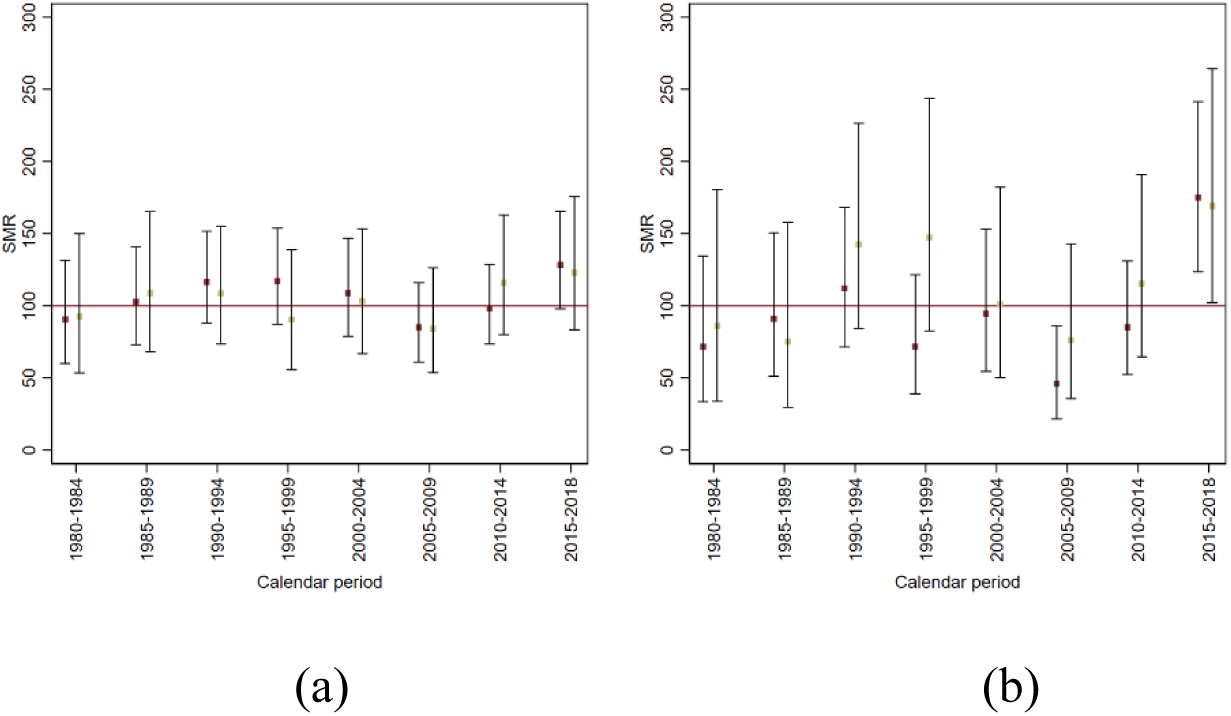
Mortality for kidney tumours. Sex-specific SMR by calendar period (Khaki Females Red Males). Panel (a) *Red area*; (b) *Red area A*. Red Area refers to the 30 Municipalities of Veneto Region connected to the PFAS contaminated water plant (aquifer), Red Area A refers to the 13 municipalities with also PFAS contamination of groundwater. 1980-2018.

**Table 5.**
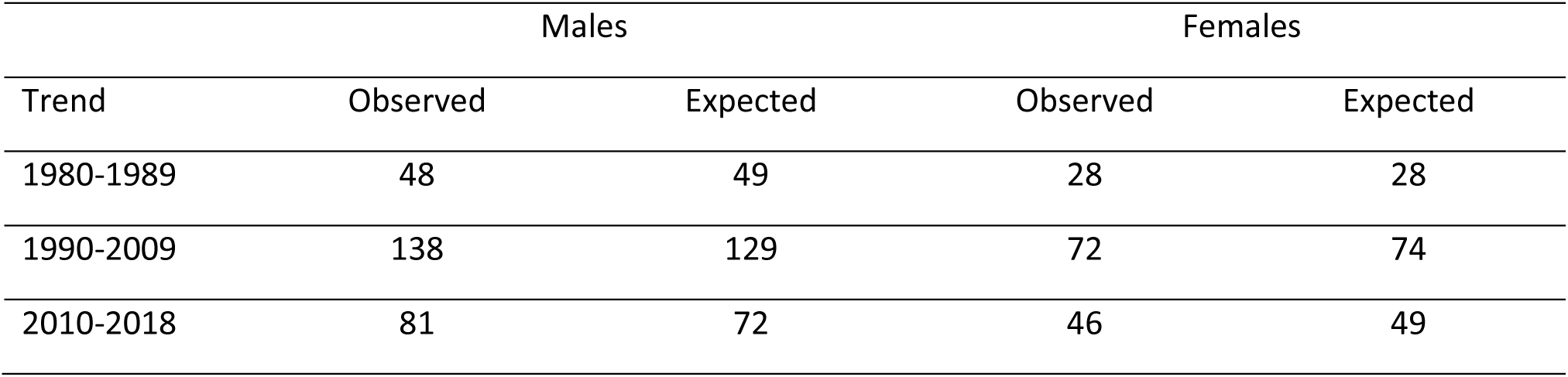
Observed and expected deaths of kidney tumours by sex and calendar periods. PFAS contaminated Red Area population. 1980-2018.

The age-sex standardized SMR for the period 2010-2018 was 1.14 (one-sided mid-p value=0.066).

In the last period observed (2015-2018), the SMR was 1.26 (90% CI: 1.02-1.54; one-sided mid-p-value=0.034) (see also Supplementary Table S5).

These values are higher for the *Red Area A*, with 41 observed deaths versus 23.7 expected (one-sided mid-p-value=0.0006). The SMR was 175 (90% CI: 124-241) for males and 169 (90% CI: 102-264) for females. The age-sex standardized SMR was 173 (90%CI: 132; 221).

We found 17 attributable cases over 41 observed in the four-year calendar period 2015-2018, corresponding to 4 attributable deaths per year in the last period of follow-up (2015-2018).

Noticeably, when restricting the analysis to renal cell kidney cancer (ICD-10 C64 code) for the period 2010-2018 in the *Red area*, we found 38 deaths versus 26 expected among males, for a SMR of 148 (90% CI: 111-194), 20 deaths versus 14 expected among females, for a SMR of 140 (90% CI: 111-204).

#### Testicular cancer mortality

In the time period 1985-1999, we observed 6 deaths versus an expected number of 4.3, for a SMR of 140 (90% supported range: 61-275) (mid-p-value=0.204). After 1999, no deaths for testicular cancer were recorded. In the first time period, 1980-1984, we observed one death versus 2.4 expected.

Restricting the analysis to the population living in the *Red Area* A, we found 5 deaths in the period 1985-1999 versus 1.95 expected, for a SMR of 256 (90% CI: 101-539; one-sided mid-pvalue= 0.032) (Supplementary Table S5).

## Discussion

### Main findings

During the 33 years from 1985 (assumed date of water contamination) to 2018 (last year of available cause-specific mortality data), in the resident population of the *Red area* (the municipalities being connect to the contaminated water plant) we observed 51,621 deaths over 47,731 expected (age-sex SMR: 108; 90% CI: 107-109). The overall burden was, therefore, an excess of 3,890 deaths (90% CI: 3543-4231). In other words, every three days we estimated 12 deaths over 11 expected deaths. We found evidence of increased mortality for diseases of the circulatory system (particularly heart diseases and ischemic heart diseases), malignant neoplastic diseases, kidney cancer and testicular cancer.

Comparing the data on the progression of contamination in time and space with the data on the production of specific molecules, it clearly appears that these effects should mainly be attributed to long-chain PFAS, that is, PFOA and PFOS [39].

### Interpretation

#### All-cause mortality

With respect to all-cause mortality, both sexes showed an excess risk. As there was strong evidence for an increase in mortality in the most recent birth cohorts, we suggest that this is the effect of PFAS exposure during human early development. An interesting finding was that, for women, the risk peaked in the birth cohort of 1945-1954, that is, for women exposed from the age of 35-39 and older. Women aged less than 35 years in 1984-1989, conversely, did not experience any excess risk. In fact, this protection status was probably accounted for by multiparous women, because nulliparous ones have been demonstrated to have higher PFAS concentrations [59]. It is worthy of note that this has also been observed in the Veneto Region, as can be seen from data in Table 3 and supplementary Table S1 in the article by Pitter et al. [34]. The protection status of multiparous women is explained by the accumulation of PFAS in the placenta [60] coupled with trans-placental transmission. This mechanism, together with breastfeeding, is the most important determinant of PFAS exposure in early life [61].

#### Circulatory disease mortality

The striking increase in mortality for diseases of the circulatory system, heart disease and ischemic heart disease is undoubtedly the most valuable finding of this study. Given the study design of our investigation, we can interpret the results as the consequences of PFAS exposure and acknowledge it as credible causal inference. The effect of PFAS exposure on cardiovascular disease is most likely mediated through the atherosclerotic process. PFOA increases the serum levels of total cholesterol and low-density cholesterol [62]. Evidence has been obtained with cross-sectional studies [22] and longitudinal studies [63]. Notably, several significant associations of PFAS serum levels with multiple markers of cardiovascular disease have also been observed in the Veneto Region, including blood pressure [64], triglyceride levels [64, 65], total cholesterol and low-density cholesterol in children and adolescents [66] as well as young adults [64].

We raise the hypothesis that a second mechanism leading to an increased risk of cardiovascular disease in PFAS-exposed populations is mediated through the occurrence of posttraumatic stress disorders caused by the severe psychologic trauma. The finding that the risk showed no or minimum decrease after 2014 provides circumstantial evidence for this. It is known that exposure to an environmental contamination resulting from industrial processes or accidents is psychologically stressful for the affected communities [67–69]. Our hypothesis is further supported by two local studies that have shown, in particular, an unfavourable impact of the contamination with PFAS in the area on the behaviour and the psychologic health of children as well as parents. Girardi et al. have shown that PFAS exposure during pregnancy and early life can adversely impact neurodevelopment and cause greater behavioral difficulties [70]. Menegatto et al. have evaluated the psychosocial impact on people who live in polluted areas and its consequences for the parental role [70, 71]. According to this second study, the uncertainty about the health effects of PFAS exposure is a major factor of stress particularly for families with children. This is due to parents’ anxiety regarding the health and the quality of life of their children, combined with the sense of responsibility and the inability to control the destiny of their families [72]. Posttraumatic stress disorders, in turn, are known to be associated with major risk factors for cardiovascular disease, including hypertension and diabetes, and with major cardiovascular disease outcomes such as myocardial infarction and heart failure [73]. In the long term, environmental stressors may lead to allostatic overload, which occurs when stress causes physiological changes and imbalances in stress mediators such as glucocorticoids, excitatory amino acids, and cytokines [74]. Through these processes, chronic stress can further increase the risk of hypertension and coronary heart disease which may also make individuals more susceptible to pollutants [75]. Chronic stress itself interacts with exposure to pollutants and amplifies their effects by compromising, for example, the immune system [76].

The importance of our study lies in the fact that, despite the evidence on atherosclerosis and posttraumatic stress disorders as direct causes of cardiovascular disease, there has been insufficient epidemiologic demonstration so far of an association between PFAS exposure and the final outcome. As has been hypothesised [77], this might be due to the anecdotally reported positive association between PFAS exposure and high-density cholesterol [66] coupled with an inverse association between the latter and the risk of cardiovascular disease. A notable detail is that, in the study area, an increased level of high-density cholesterol in people exposed to PFAS has actually been demonstrated [64, 66]. Apart from this, the role of unknown confounders in masking the indirect relationship between PFAS exposure and cardiovascular disease has been discussed [78]. Our study, which took advantage of the natural experiment in the *Red area*, and, as a consequence, the present study is the first to indicate a clear role of PFAS exposure on cardiovascular mortality.

It is also necessary to draw attention to the consistency between our results about mortality from circulatory diseases and the ecological study by Mastrantonio et al., conducted in the same geographic area, given the differences in design, identification of the exposed population and potentially associated biases [79].

In the *Red area*, incidentally, directly standardized (2013 European standard population) mortality rates for circulatory system disease in 1985-2018, i.e. 35.9 per 10,000 among men and 26.5 among women, exceed those observed in Italy as a whole in 2019, i.e. 33.3 and 23.7, respectively (data not shown).

#### Kidney cancer and testicular cancer mortality

The International Agency for Research on Cancer has recently classified PFOA as ‘carcinogenic to humans’ (Group 1) and PFAS as ‘possibly carcinogenic to humans’ (Group 2B) [28]. With respect to single cancer sites, there is ‘limited’ epidemiologic evidence in humans for an association between PFOA exposure and kidney cancer as well as testicular cancer. Our findings add some evidence to the existing literature. As far as kidney cancer is concerned, we found a consistent increase in mortality in the exposed population and even higher in the *Red area* A population. Steenland and Woskie observed a risk increase in a cohort of subjects occupationally exposed to PFOA [31], which was not the case for the occupational cohort reported by Raleigh et al. [80]. In a previous ecologic mortality study from the Veneto Region comparing areas with and without PFOA-contaminated drinking water, a moderate excess risk of kidney cancer among women (SMR: 1.32; 95%, CI: 1.06–1.65) was found [79]. In a case-control study, a positive exposure–response trend for several PFAS including PFOA was observed [81]. These findings should be considered in relation to the fact that tetrafluoroethylene is a kidney carcinogen in rodents [82] and that exposure to PFOA and exposure to tetrafluoroethylene are strongly correlated [31]. No animal studies have found an association between PFOA and kidney cancer, but PFOA concentrates in renal tissues. This means that our finding is plausible.

With respect to testicular cancer, mortality is a weak outcome due to the recent marked improvement in survival. However, before year 2000, we found a consistent increase in mortality, particularly in the most exposed population living in *Red Area A*. For the period 1997-2014, an analysis on hospital discharge data on orchiectomies for testicular cancer showed a positive association with PFOA exposure at the ecological level [28]. Furthermore, an ecologic study has already found higher mortality in PFAS-exposed areas of the Veneto Region [79]. An excess risk was reported by a cohort study with high exposure to PFOA [83] and a case-control study [32]. Both found a clear positive exposure-response. PFOA exposure is associated with testicular cancer in rodents [29]. According to a literature review, the epidemiologic evidence is supportive but not definitive [77]. Caution was also suggested by a second review [29], because the mechanism by which PFOA causes tumours in rats seems less relevant in humans [84]. According to a meta-analysis, conversely, the link with testicular cancer is probably a causal one [85]. In part, this uncertainty reflects methodological constraints. Case ascertainment in the evaluation of the risk of testicular cancer is almost certainly biased downward if the endpoint is mortality, because cisplatin-based combination chemotherapy and the refinement of post-chemotherapy surgical procedures have greatly improved long-term survival in most patients [86].

### Comparison with previous research from the area of contamination

Several previous studies have evaluated the health surveillance data collected in the *Red area*. The degree of consistency between their results and our own is considerable. Statistically significant associations of PFAS serum levels with blood pressure and triglyceride levels, two components of the metabolic syndrome, have been observed [65]. In children and adolescents, significant associations have been detected between PFOA, PFOS, PFHxS, PFNA, and tPFAS and total cholesterol, low-density lipoprotein cholesterol and, to a lesser extent, high-density lipoprotein cholesterol [66]. In young adults, PFOA, PFOS, and PFHxS were associated with total cholesterol, low-density lipoprotein cholesterol and high-density lipoprotein cholesterol, with PFOA and PFHxS being also associated with triglycerides [64]. An association between the serum levels of PFOA, PFOS, PFHxS and PFNA and multiple markers of cardiovascular risk was also found in a subgroup of 232 men formerly employed in the factory of Trissino and attending the health surveillance programme [87]. A possible influence of PFAS exposure on lipid metabolisms during pregnancy has been reported [88].

An ecological mortality study showed higher rates for multiple causes of death in the contaminated municipalities [79]. An occupational cohort study has been conducted on workers employed in the factory of Trissino. The study found an increase of the risk of liver cancer, liver cirrhosis, diabetes, and haematologic malignancies in 462 men in association with the highest cumulative internal dose of PFOA [39]. Others have suggested an additional causal role of PFAS in the atherosclerosis process, mediated through an impaired downstream signaling of platelets’ activation and aggregation [89].

### Methodological issues

Some methodological issues of this study need to be mentioned. First, the study is based on aggregate data at the municipality level and the interpretation of the results depends on the assumption of homogeneity of exposure within municipalities – which is almost certain for the *Red Area A* population. The choice of using the population of the provinces of Vicenza, Verona, and Padova as reference could bias conservatively our results. However, a sensitivity analysis conducted excluding the exposed population from the reference gave similar results. In addition, all time period comparisons were done taking as reference the time periods before the relevant exposure happened. This internal analysis reassures in terms of comparability of exposed and unexposed, and the use of SMR guarantees that confounding by secular trend in the time evolution of mortality is controlled for.

Second, we could not adjust the association of PFAS exposure with the risk of cardiovascular disease for those conditions acting as mediators of the effect of PFAS, for example low-density cholesterol and hypertension. It must be noted that, otherwise, we would have eclipsed the effect of PFAS, which depends on mediating factors. It may be that this is an additional explanation for the frequent failures of the epidemiologic research on PFAS-associated cardiovascular disease.

Third, our observation did not start at the time of first exposure but approximately 15 years later because of limitations in availability of mortality data and, moreover, we considered the Red Area population as exposed, excluding those municipalities with groundwater contamination since 1966. People exposed earlier may have died before the start of the study and, if so, their death would not be documented. In particular, people most susceptible to the effects of exposure may have developed the disease earlier. These factors may have led to an underestimate of the risk.

And fourth, comparisons with other studies from the same area should be made with some caution, because of the possibility that the municipalities considered to represent the ‘contaminated area’ and the uncontaminated (or reference) area have been selected using data from different reports of regional authorities or have been defined according to criteria partly different from ours [79]. The selection of the uncontaminated area is particularly prone to arbitrary assumptions and between-studies variation, with a risk of misclassification of exposure. We took the reference municipalities within the boundaries of the three provinces involved. They probably had some, albeit low, degree of contamination with PFAS but they were much more similar to the contaminated municipalities for many potential confounders.

### Future perspectives

Global concerns about the public health impact of PFAS are growing and the scientific, political and social attitudes towards the PFAS problem are evolving. The revision of the IARC classification of PFOA to “carcinogenic to humans” (Group 1) and PFOS to ‘possibly carcinogenic to humans’ (Group 2B) [28] will most likely boost this process. In the first place, the transition to the replacements of PFAS and the creation of comprehensive environmental monitoring programmes will become important topics in many agendas. The combination of regulative restrictions and pan-European biomonitoring programmes is considered the key strategy for PFAS control across the EU [27]. Increasing attention is also being paid to the creation of groundwater parks, that is, areas free of agricultural activities and nutritious sewage sludge.

Although effective strategies for removing PFAS do exist [8], research efforts on technologies and water treatment systems should continue [90]. PFAS treatment and remediation costs, too, have become a dominant topic of environmental policies and among regulators and utilities, and the pressure to find effective and economical solutions is increasing [91, 92].

The exploration of options for PFAS control plans is paralleled by research efforts and advancements on the mechanisms of carcinogenicity. An emerging view is that there are distinct but interdependent general pathways of PFAS action involving, in particular, metabolism, endocrine disruption and epigenetic perturbation [93]. These three pathways seem mutually enforcing and may combine to establish a pro-tumorigenic environment. Also, these pathways are predicted to be dependent on the dose and the window of exposure during life. This might contribute to explain the difficulties in obtaining epidemiologic and experimental evidence linking PFAS and cancer. The relationship between prenatal and postnatal exposure to PFAS, inflammation status and cardiometabolic factors is another promising area of research [94].

As a final remark, concerning health surveillance programmes, our findings support the view that more consideration should be given to the psychological impact of environmental pollution, which is poorly recognized by the health authorities responsible for managing disasters [71].

## Conclusions

For the first time, the association of PFAS with mortality from cardiovascular disease was formally demonstrated in the world’s largest exposed population. Moreover, circumstantial evidence regarding kidney cancer and testicular cancer is consistent with previously reported data. Given the present results and the recent IARC revision, it is urged to have an immediate ban of PFAS production and to start implementing additional remediation activities in contaminated areas.

## Declarations

### Ethics approval and consent to participate

The study was approved by the Ethics Committee at the Romagna Cancer Institute (ID: IRST100.37).

### Consent for publication

Not applicable.

### Data availability

Research data are available from Giada Minelli (contact) upon reasonable request.

### Competing interests

The authors declare no competing interests.

### Funding

This study was partially funded by the Veneto Regional Administration (DGR 1495/2019, DDR 30/2020, Resolution AULSS8 1402/2020) by committing a professional assignment to the Giulio A. Maccacaro Epidemiology & Prevention Society. The authors received no funding. The authors retained full control over the manuscript and full intellectual freedom to report and interpret the results.

---

**Supplementary Figure S1.**
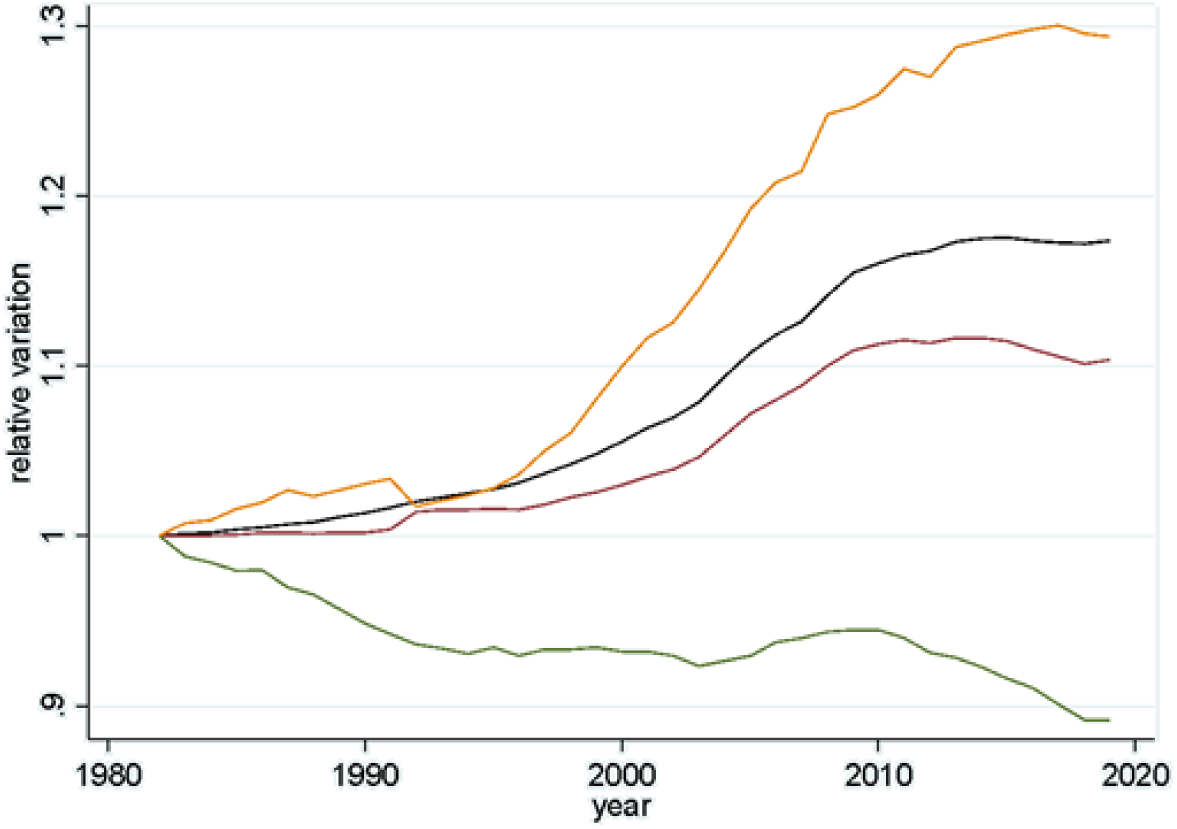
Relative variation of population size (reference calendar year 1982). Three Provinces of Vicenza, Verona and Padova (black line), the 30 Municipalities of the PFAS contaminated Red Area (red line), the municipality of Lonigo (yellow line) and Montagnana (green line).

**Supplementary Figure S2.**
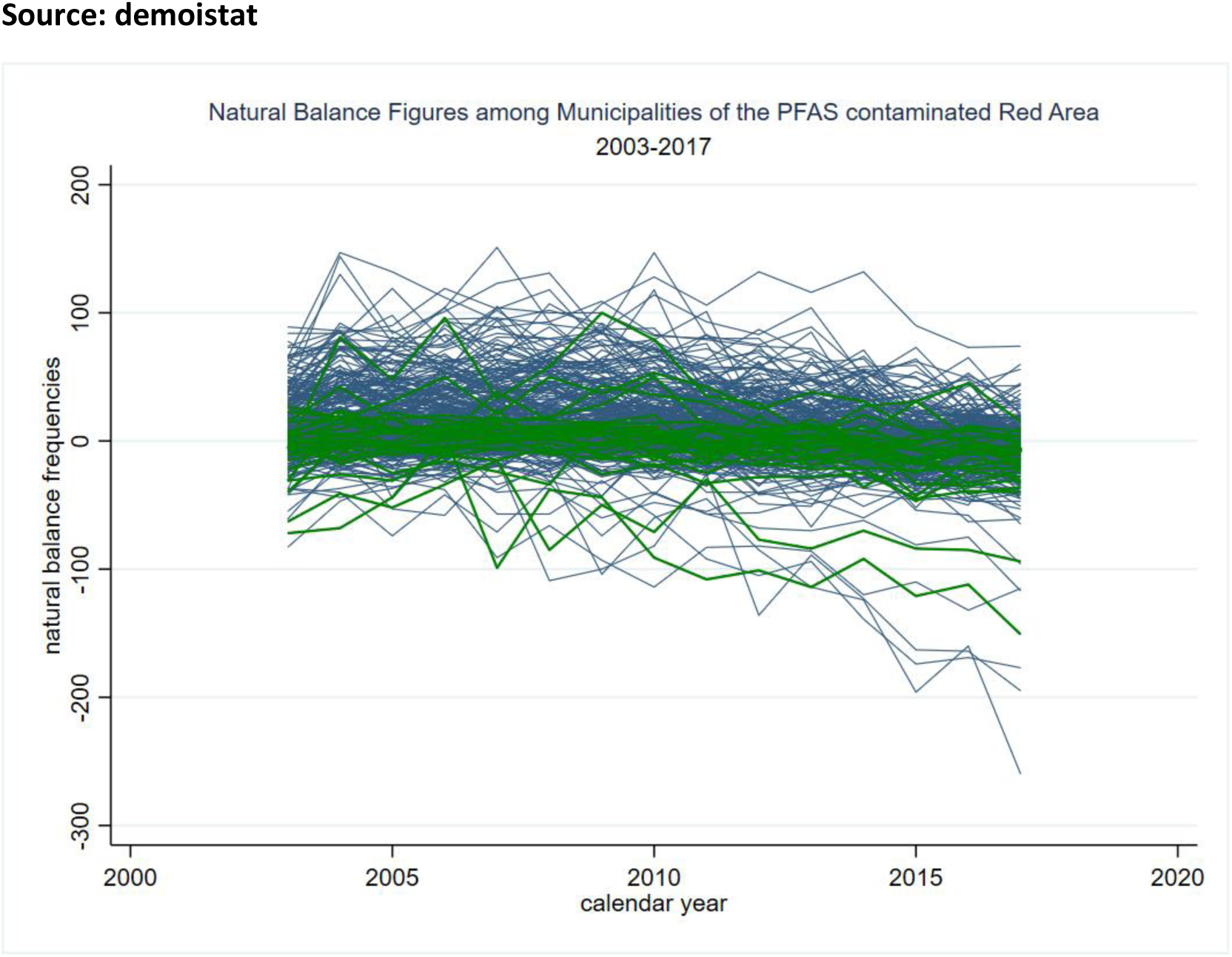
Natural balance by calendar year for 321 municipalities of the Provinces of Vicenza, Verona and Padova. (Green lines: the 30 Municipalities of the PFAS contaminated Red Area). The capital cities were excluded. The The municipalities of the Red Area with lowest natural balance were Legnago and Montagnana, the municipality with positive balance was Lonigo.

**Supplementary Figure S3.**
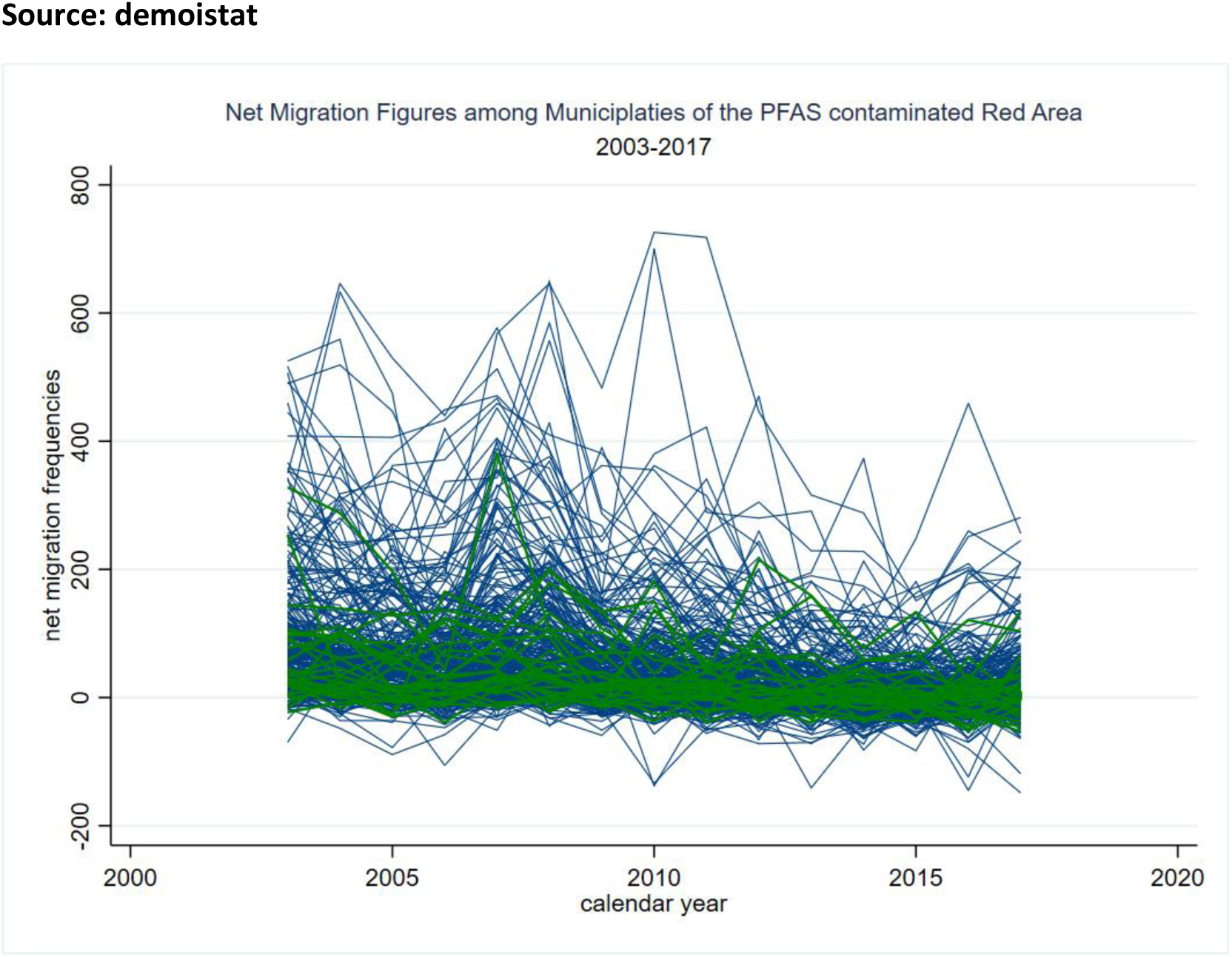
Net migration by calendar year for 321 municipalities of the Provinces of Vicenza, Verona and Padova. (Green lines: the 30 Municipalities of the PFAS contaminated Red Area). The capital cities were excluded. The municipality of the Red Area with highest net migration was the municipality of Lonigo.

**Supplementary Figure S4.**
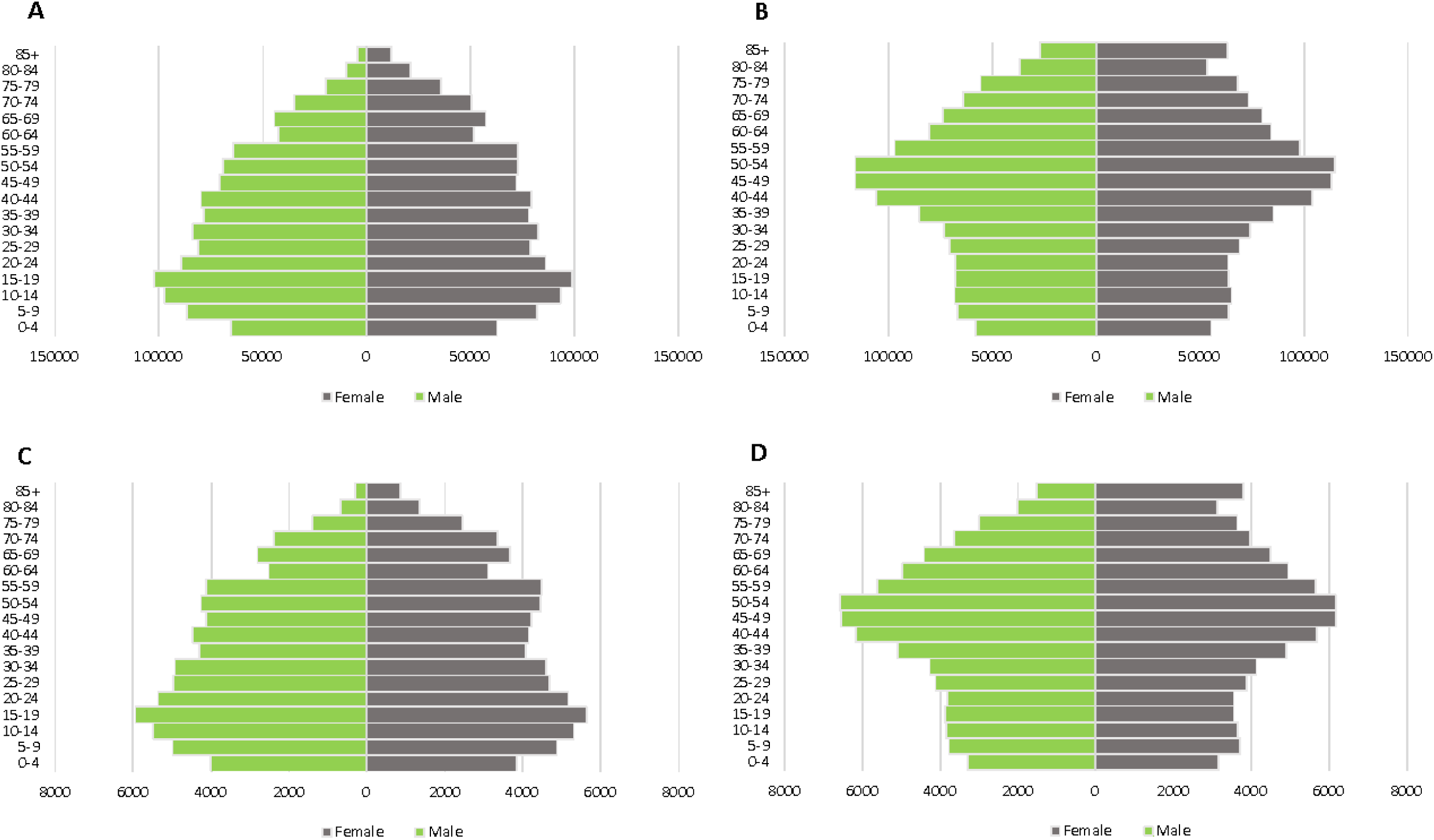
Resident population age-sex pyramid of the Provinces of Vicenza, Verona and Padova (panel A and B) and the 30 Municipalities of the PFAS contaminated Red Area (panel C and D). 1^st^ January 1982 (panel A and C) and 1^st^ January 2018 (panel B and D). Source: Italian National Institute of Statistics (ISTAT)

**Supplementary Figure S5.**
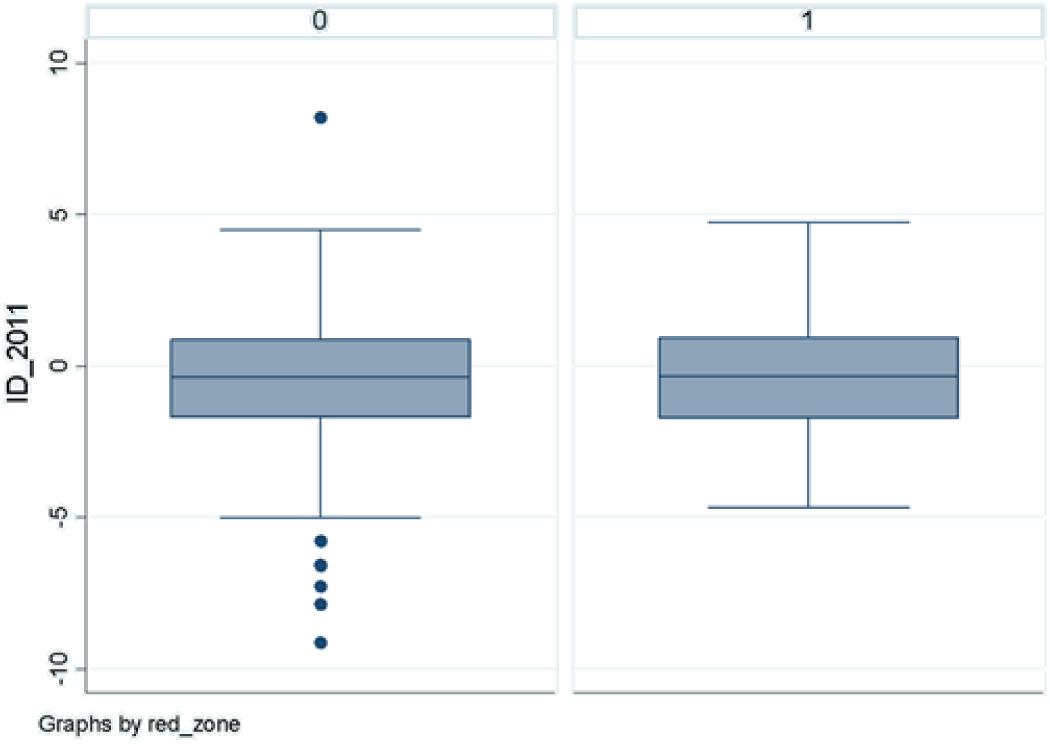
Box and whiskers plots of material deprivation index at 2011 census for the three provinces of Vicenza, Verona and Padova, and for the 30 Municipalities of the PFAS contaminated Red Area. Source: Rosano A, Pacelli B, Zengarini N, Costa G, Cislaghi C, Caranci N. Aggiornamento e revisione dell’indice di deprivazione italiano 2011 a livello di sezione di censimento [Update and review of the 2011 Italian deprivation index calculated at the census section level]. Epidemiol Prev. 2020 Mar-Jun;44(2-3):162-170. Italian. doi: 10.19191/EP20.2-3.P162.039

**Supplementary Figure S6.**
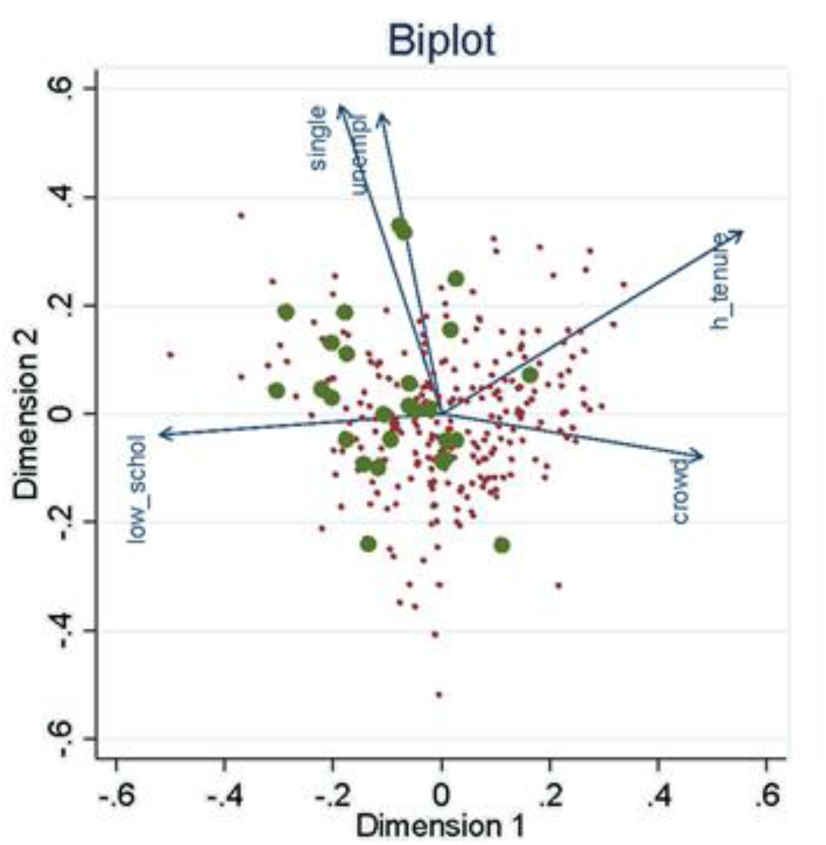
Biplot of the first two principal components of five socio-demographics indicators (low scholarity, unemployment, house crowding, house tenure and single parenting). Points represent the 321 municipalities of the three provinces of Vicenza, Verona and Padova. In green the 30 Municipalities of the PFAS contaminated Red Area. **Source: Rosano A, Pacelli B, Zengarini N, Costa G, Cislaghi C, Caranci N. Aggiornamento e revisione dell’indice di deprivazione italiano** 2011 **a livello di sezione di censimento [Update and review of the** 2011 **Italian deprivation index calculated at the census section level]. Epidemiol Prev.** 2020 **Mar-Jun;44(2-3):162-170. Italian.** doi: 10.19191/EP20.2-3.P162.039

**Supplementary Figure S7.**
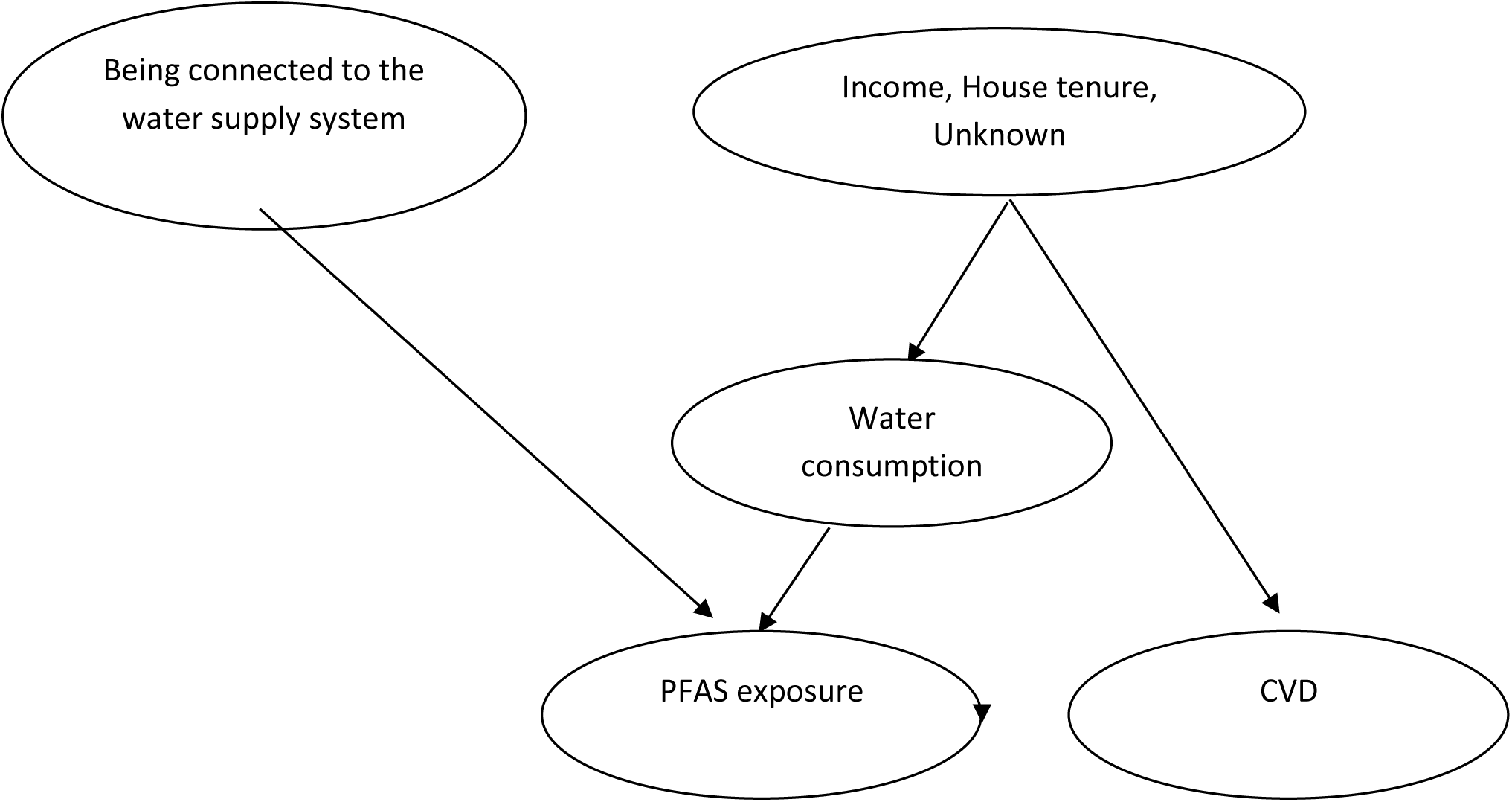
Directed acyclic graph of the assumed causal model.

**Supplementary Table S1.** Distribution of perfluoroctanic acid (PFOA) serum concentrations (ng/ml) by attained at first invitation and year of invitation. Veneto Region Health Surveillance. In each cell of the table number of subjects, 5° percentile, mean (in bold), median, 95° percentile. When the number of subjects <50 the cell is left empty. P= <14 yrs old at invitation. Source: Regione del Veneto. Piano di sorveglianza sanitaria sulla popolazione esposta a PFAS. Rapporto n. 19 Ottobre 2023.

| Age class<br>(years) | Year of invitation |  |  |
| --- | --- | --- | --- |
|  | 2017-2018 | 2019-2020 | 2021-2023 |
| **58-72** | 0 | 3480 | 7263 |
|  | 0 | 9.8 | 3.2 |
|  | **0** | **111.3** | **81.9** |
|  | 0 | 78.5 | 50.4 |
|  | 0 | 309.8 | 265.3 |
| **45-61** | 1472 | 8443 | 1771 |
|  | 9.1 | 3.6 | 3.1 |
|  | **91.8** | **71.2** | **58.9** |
|  | 64.7 | 41.1 | 34.3 |
|  | 264.7 | 240.7 | 187 |
| **36-51** | 7042 | 4841 | 78 |
|  | 3.6 | 2.3 | 1.2 |
|  | **62** | **42.8** | **26.6** |
|  | 33.7 | 22.1 | 12.9 |
|  | 217.7 | 149 | 112.2 |
| **26-41** | 7783 | 847 | 100 |
|  | 3.3 | 1.5 | 0.4 |
|  | **60.5** | **31.4** | **20.8** |
|  | 34.8 | 15.1 | 8.5 |
|  | 203.3 | 118.7 | 93.8 |
| **15-31** | 8721 | 899 | 263 |
|  | 7.2 | 2.5 | 0.8 |
|  | **65.2** | **28.6** | **15.9** |
|  | 50.6 | 20.1 | 9.8 |
|  | 170.3 | 81.4 | 48.5 |
| **15-17** | 0 | 1848 | 0 |
|  | 0 | 4.2 | 0 |
|  | **0** | **28.7** | **0** |
|  | 0 | 24.2 | 0 |
|  | 0 | 67 | 0 |
| **14-18** | 0 | 96 | 1167 |
|  | 0 | 2.2 | 1.6 |
|  | **0** | **21.9** | **12.9** |
|  | 0 | 19 | 9.9 |
|  | 0 | 52 | 33 |
| **14-16** | 0 | 0 | 660 |
|  | 0 | 0 | 1.7 |
|  | **0** | **0** | **9.8** |
|  | 0 | 0 | 7.9 |
|  | 0 | 0 | 22.4 |
| **8-12 (P)** | 182 | 1713 | 0 |
|  | 7.7 | 4.6 | 0 |
|  | **30.7** | **26.8** | **0** |
|  | 25.8 | 21.7 | 0 |
|  | 69.1 | 64.4 | 0 |
| **8-13 (P)** | 0 | 873 | 552 |
|  | 0 | 3.9 | 1.2 |
|  | **0** | **23.6** | **7.8** |
|  | 0 | 17.9 | 5.6 |
|  | 0 | 64.2 | 23.6 |
| **8-11 (P)** | 0 | 0 | 746 |
|  | 0 | 0 | 1.1 |
|  | **0** | **0** | **7.8** |
|  | 0 | 0 | 5 |
|  | 0 | 0 | 22.5 |
| **7-9 (P)** | 0 | 0 | 257 |
|  | 0 | 0 | 1.2 |
|  | 0 | 0 | 8.2 |
|  | 0 | 0 | 4.9 |
|  | 0 | 0 | 24.4 |

**Supplementary Table S2.** Distribution of perfluorosulfonic acid (PFOS) serum concentrations (ng/ml) by attained at first invitation and year of invitation. Veneto Region Health Surveillance. In each cell of the table number of subjects, 5° percentile, mean (in bold), median, 95° percentile. When number of subject <50 the celli is leave empty. P= <14 yrs old at invitation. Source: Regione del Veneto. Piano di sorveglianza sanitaria sulla popolazione esposta a PFAS. <u>Rapporto n.</u> <u>19 Ottobre 2023.</u>

| Age class<br>(years) | Year of invitation |  |  |
| --- | --- | --- | --- |
|  | 2017-2018 | 2019-2020 | 2021-2023 |
| **58-72** | 0 | 3480 | 7263 |
|  | 0 | 2 | 1.6 |
|  | **0** | **6.2** | **6.6** |
|  | 0 | 5.3 | 5.3 |
|  | 0 | 13.1 | 15.3 |
| **45-61** | 1472 | 8443 | 1771 |
|  | 1.6 | 1.4 | 1.4 |
|  | **5.5** | **5.8** | **5.2** |
|  | 4.7 | 4.6 | 4.1 |
|  | 12 | 13.8 | 12.1 |
| **36-51** | 7042 | 4841 | 78 |
|  | 1.2 | 1.3 | 0.7 |
|  | **4.8** | **4.8** | **4.3** |
|  | 3.8 | 3.7 | 3.5 |
|  | 11 | 11.4 | 10.2 |
| **26-41** | 7783 | 847 | 100 |
|  | 1.3 | 1.2 | 0.6 |
|  | **4.7** | **3.8** | **3** |
|  | 3.8 | 3 | 2.3 |
|  | 10.5 | 9.3 | 7.6 |
| **15-31** | 8721 | 899 | 263 |
|  | 1.5 | 1.1 | 0.6 |
|  | **4.7** | **3.3** | **2.9** |
|  | 3.8 | 2.7 | 2.5 |
|  | 10.3 | 7.2 | 6.1 |
| **15-17** | 0 | 1848 | 0 |
|  | 0 | 1.1 | 0 |
|  | **0** | **2.9** | **0** |
|  | 0 | 2.5 | 0 |
|  | 0 | 6.2 | 0 |
| **14-18** | 0 | 96 | 1167 |
|  | 0 | 1 | 0.7 |
|  | **0** | **2.1** | **2.1** |
|  | 0 | 1.8 | 1.8 |
|  | 0 | 4.3 | 4.5 |
| **14-16** | 0 | 0 | 660 |
|  | 0 | 0 | 0.7 |
|  | **0** | **0** | **2.1** |
|  | 0 | 0 | 1.7 |
|  | 0 | 0 | 4.4 |
| **8-12 (P)** | 182 | 1713 | 0 |
|  | 1.1 | 0.9 | 0 |
|  | **3.3** | **2.4** | **0** |
|  | 2.4 | 2 | 0 |
|  | 5.4 | 4.9 | 0 |
| **8-13 (P)** | 0 | 873 | 552 |
|  | 0 | 1.1 | 0.4 |
|  | **0** | **2.8** | **1.6** |
|  | 0 | 2.4 | 1.3 |
|  | 0 | 5.6 | 3.7 |
| **8-11 (P)** | 0 | 0 | 746 |
|  | 0 | 0 | 0.5 |
|  | **0** | **0** | **1.7** |
|  | 0 | 0 | 1.4 |
|  | 0 | 0 | 4 |
| **7-9 (P)** | 0 | 0 | 257 |
|  | 0 | 0 | 0.5 |
|  | **0** | **0** | **1.6** |
|  | 0 | 0 | 1.2 |
|  | 0 | 0 | 4.2 |

**Supplementary Table S3.** Summary of the ICD9 and ICD10 version 2019 codes used in the study with details regarding the causes of death considered in the study.

| ICD9 | ICD10 | Category | Details |
| --- | --- | --- | --- |
| 001-999 | A00-Z99.9 | All causes |  |
| 140-208 | C00-C97 | Malignant neoplasms | All malignant neoplasms, including the categories analysed in detail in this analysis |
| 155 | C22 | Malignant neoplasm of liver and intrahepatic bile ducts | Malignant neoplasm of liver and intrahepatic bile ducts, excluding gallbladder and extrahepatic bile ducts |
| 155.0 | C22.0, C22.2 | Malignant neoplasm of liver | Malignant neoplasm of liver including liver cell carcinoma and hepatoblastoma |
| 157 | C25 | Malignant neoplasm of pancreas | Malignant neoplasm of pancreas |
| 162 | C33-C34 | Malignant neoplasm of trachea, bronchus and lung | Malignant neoplasm of trachea, bronchus and lung excluding pleura |
| 174 | C50 | Malignant neoplasm of breast | Malignant neoplasms of female breast |
| 182 | C54 | Malignant neoplasm of corpus uteri | Malignant neoplasm of corpus uteri |
| 183 | C56-C57 | Malignant neoplasms of ovary and other and unspecified female genital organs | Malignant neoplasms of ovary and other and unspecified female genital organs. They include Fallopian tube, broad ligament, round ligament and parametrium. |
| 186 | C62 | Malignant neoplasm of testis | Malignant neoplasm of testis. They include malignant tumor of undescended, descended and unspecified testis |
| 189 | C64-C66, C68 | Malignant neoplasms of kidney and other and unspecified urinary organs | Malignant neoplasm of kidney, renal pelvis, ureter, urethra, paraurethral glands and contiguous or overlapping sites, excluding bladder |
| 193 | C73 | Malignant neoplasm of thyroid gland | Malignant neoplasms of thyroid gland |
| 200-208 | C81-C96 | Malignant neoplasms of lymphoid, haematopoietic and related tissue | All solid and non-solid malignant tumors of the lymphatic and hematopoietic tissue |
| 250 | E10-E14 | Diabetes mellitus | Diabetes mellitus including acute problems (e.g. hypoglycaemic coma, ketoacidosis) and complications (renal or ophthalmic complications, neurological and peripheral circulatory complications) |
| 390-459 | I00-I99 | Diseases of the circulatory system | Including the categories of heart disease described below, cerebrovascular disease, diseases of the arteries (atherosclerosis, aneurysms, embolisms and vasculitis), veins (thrombosis and embolisms) and lymphatic vessels |
| 390-429 | I00-I09, I11, I13, I20-I51 | Heart diseases | Including rheumatic, hypertensive, and ischaemic heart diseases described below, pulmonary circulation diseases, pericardial, endocardial and myocardial disease, conduction disorders and heart failure |
| 410-414 | I20-I25 | Ischaemic heart diseases | Ischaemic heart diseases |
| 520-579 | K00-K93 | Diseases of the digestive system | Diseases of the digestive system |

**Supplementary Table S4.** Observed deaths, directly standardized mortality rates (2013 European standard population) per 100,000, 0-59 and 60-85 cumulative mortality rates, attributable deaths and probability of 0 attributable deaths by causes of death, area, sex and calendar period. Red Area refers to the 30 Municipalities of Veneto Region connected to the PFAS contaminated water plant (aquifer), Red Area A refers to the 13 municipalities with also PFAS contamination of groundwater. 1980-2018. **See file “Supplementary Table S4.xlsx”**

**Supplementary Table S5.**
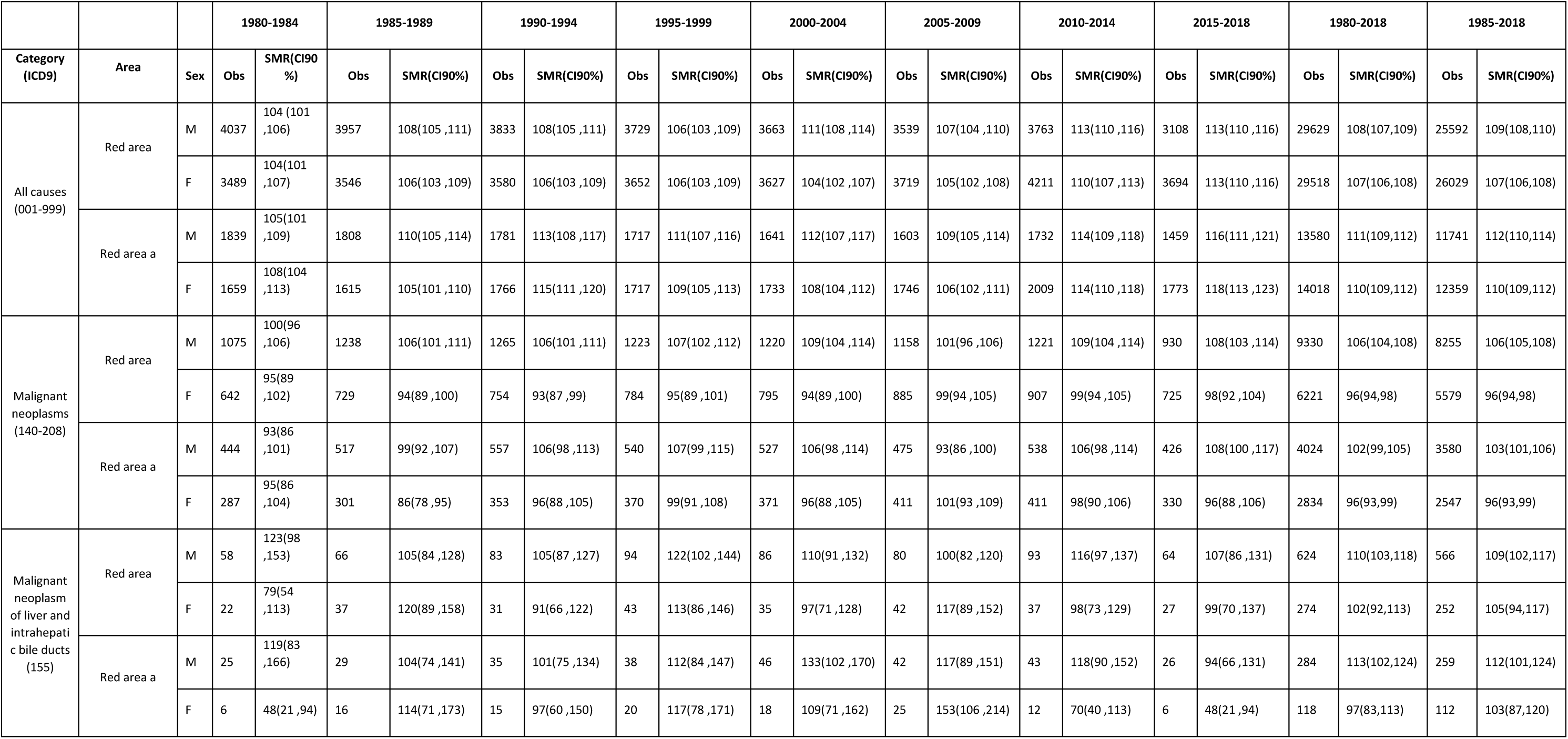
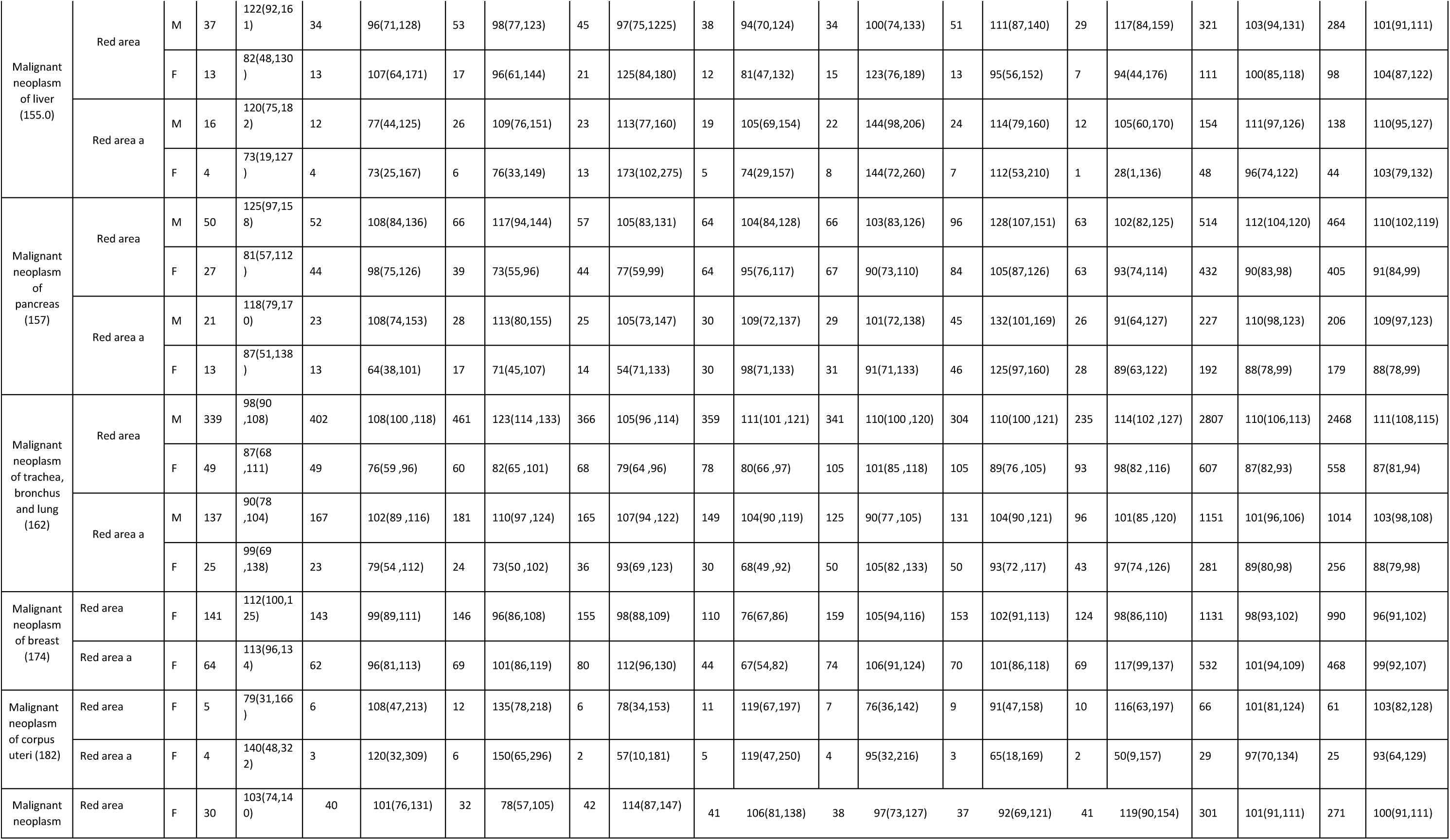
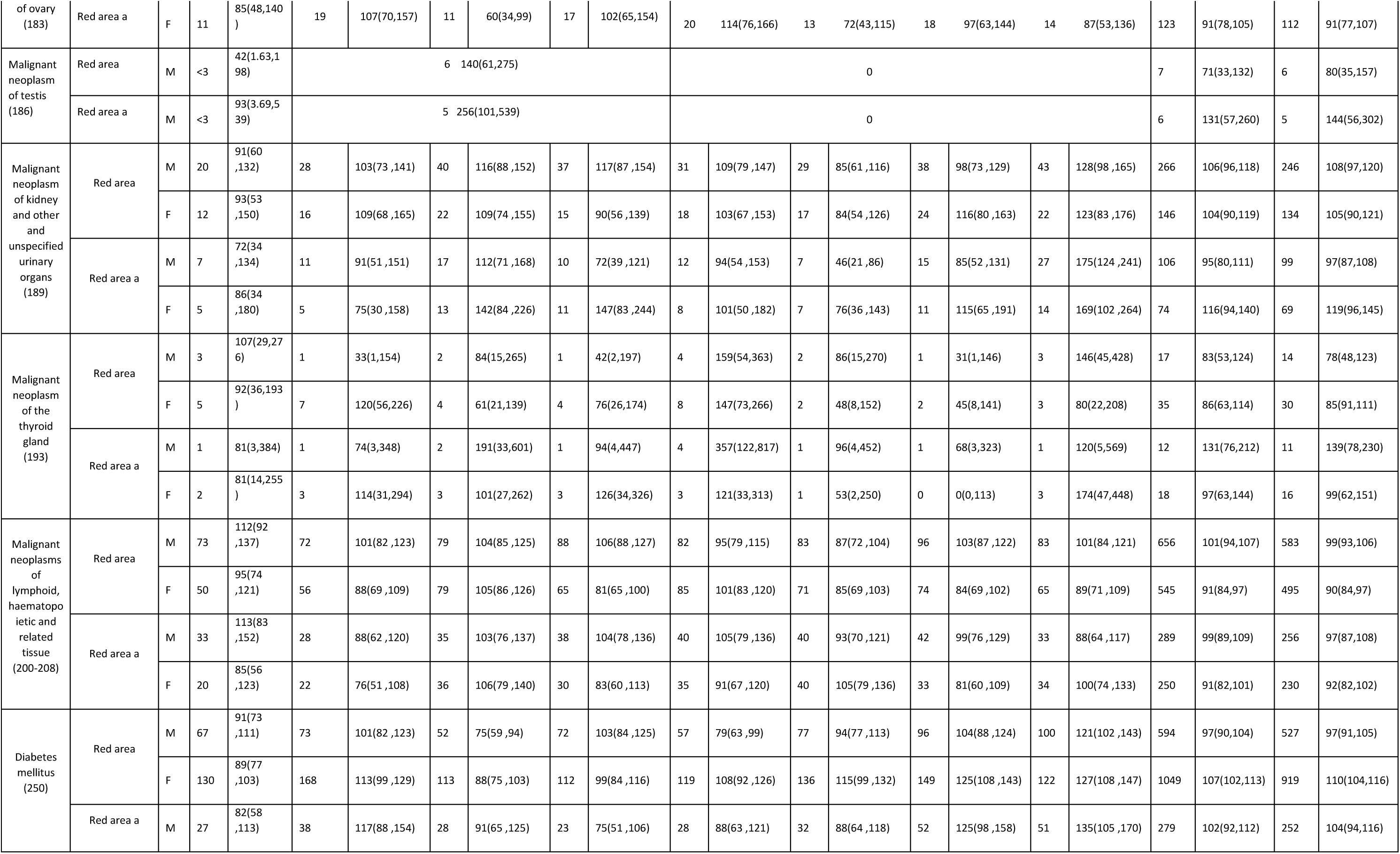
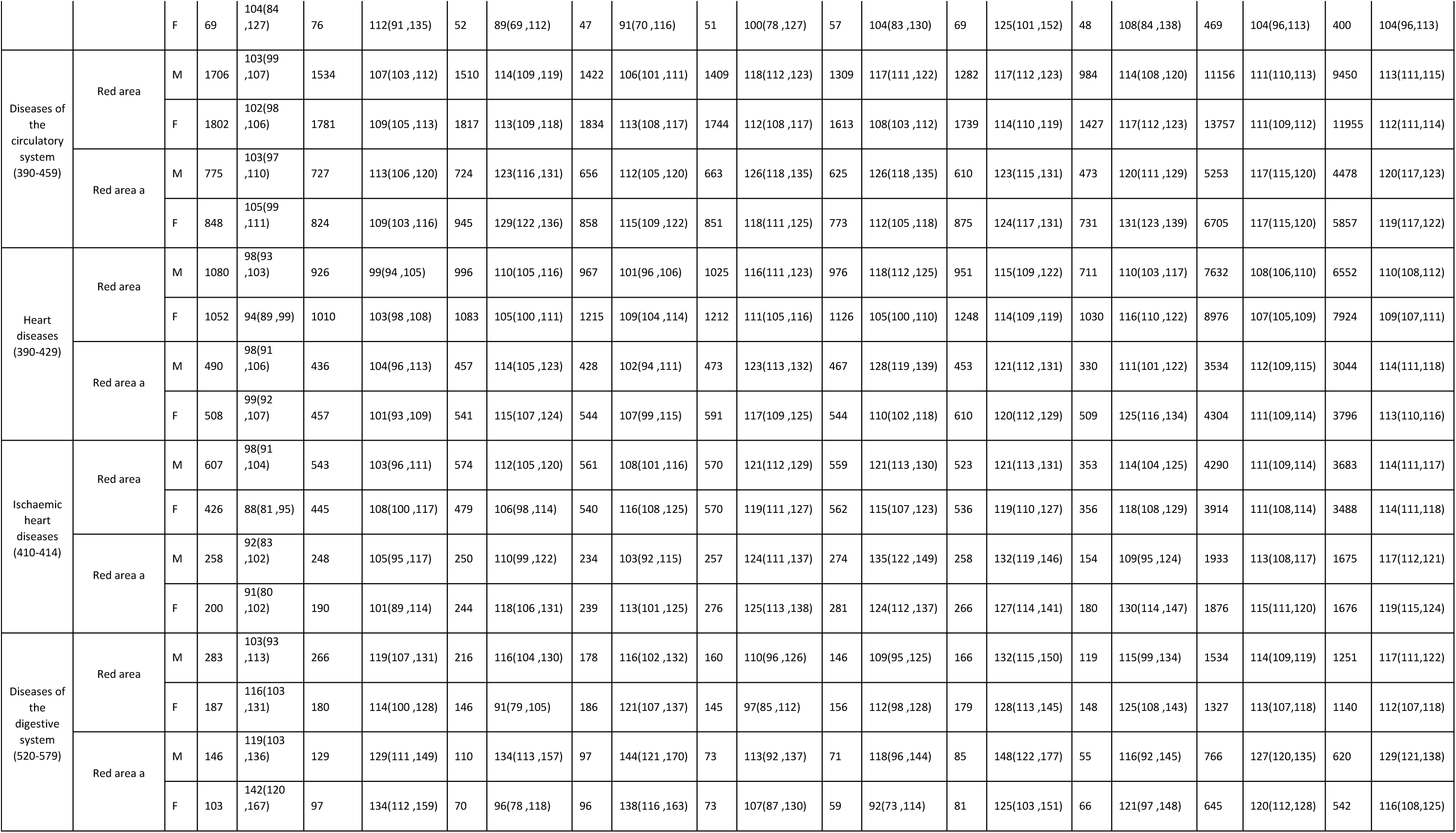
Observed number of deaths, Standardized mortality ratios and 90% Confidence Interval (CI) by causes of death, area, sex and calendar period. Red Area refers to the 30 Municipalities of Veneto Region connected to the PFAS contaminated water plant (aquifer), Red Area A refers to the 13 municipalities with also PFAS contamination of groundwater. 1980-2018.

**Supplementary Table S6.** Observed number of deaths, Standardized mortality ratios, ratio of SMRs (rr), exact onesided midpvalues by causes of death, area, sex and broad calendar period (1980-2018, 1980-1989, 2010-2018). Red Area refers to the 30 Municipalities of Veneto Region connected to the PFAS contaminated water plant (aquifer), Red Area A refers to the 13 municipalities with also PFAS contamination of groundwater.

| obs_80_18 | smr_80_18 | obs_80_89 | smr_80_89 | obs_10_18 | smr_10_18 | rr_start | rr_end | midp_start | midp_end | categoryicd9 |
| --- | --- | --- | --- | --- | --- | --- | --- | --- | --- | --- |
| 29629 | 108 | 7994 | 106 | 6871 | 113 | 0.98 | 1.05 | 0.05 | 0 | All causes (001-999) |
| 29518 | 107 | 7035 | 105 | 7905 | 111 | 0.98 | 1.04 | 0.06 | 0 |  |
| 13580 | 111 | 3647 | 107 | 3191 | 115 | 0.96 | 1.04 | 0.01 | 0.02 |  |
| 14018 | 110 | 3274 | 106 | 3782 | 116 | 0.96 | 1.05 | 0.02 | 0 |  |
| 9330 | 106 | 2313 | 103 | 2151 | 109 | 0.97 | 1.03 | 0.08 | 0.1 | Malignant neoplasms (140-208) |
| 6221 | 96 | 1371 | 94 | 1632 | 99 | 0.98 | 1.03 | 0.22 | 0.11 |  |
| 4024 | 102 | 961 | 96 | 964 | 107 | 0.94 | 1.05 | 0.03 | 0.07 |  |
| 2834 | 96 | 588 | 90 | 741 | 97 | 0.94 | 1.01 | 0.06 | 0.39 |  |
| 624 | 110 | 124 | 113 | 157 | 112 | 1.03 | 1.02 | 0.62 | 0.41 | Malignant neoplasm of liver and intrahepatic bile ducts (155) |
| 274 | 102 | 59 | 101 | 64 | 98 | 0.99 | 0.96 | 0.48 | 0.62 |  |
| 284 | 113 | 54 | 110 | 69 | 108 | 0.97 | 0.96 | 0.43 | 0.64 |  |
| 118 | 97 | 22 | 83 | 18 | 61 | 0.86 | 0.63 | 0.24 | 0.98 |  |
| 321 | 103 | 71 | 108 | 80 | 113 | 1.05 | 1.10 | 0.66 | 0.20 | Malignant neoplasm of liver and intrahepatic bile ducts (155.0) |
| 111 | 100 | 26 | 93 | 20 | 95 | 0.93 | 0.95 | 0.37 | 0.58 |  |
| 154 | 111 | 28 | 97 | 36 | 111 | 0.87 | 1.00 | 0.24 | 0.49 |  |
| 48 | 96 | 8 | 63 | 8 | 82 | 0.66 | 0.85 | 0.11 | 0.65 |  |
| 514 | 112 | 102 | 116 | 159 | 116 | 1.04 | 1.04 | 0.66 | 0.31 | Malignant neoplasm of pancreas (157) |
| 432 | 90 | 71 | 78 | 147 | 99 | 0.87 | 1.10 | 0.10 | 0.12 |  |
| 227 | 110 | 44 | 113 | 71 | 113 | 1.03 | 1.03 | 0.59 | 0.39 |  |
| 192 | 88 | 26 | 73 | 74 | 109 | 0.83 | 1.24 | 0.17 | 0.04 |  |
| 2807 | 110 | 741 | 103 | 539 | 112 | 0.94 | 1.02 | 0.04 | 0.34 | Malignant neoplasm of trachea, bronchus and lung (162) |
| 607 | 87 | 98 | 81 | 198 | 93 | 0.93 | 1.07 | 0.24 | 0.17 |  |
| 1151 | 101 | 304 | 96 | 227 | 103 | 0.95 | 1.02 | 0.19 | 0.38 |  |
| 281 | 89 | 48 | 88 | 93 | 95 | 0.99 | 1.07 | 0.48 | 0.26 |  |
| 1131 | 98 | 284 | 105 | 277 | 100 | 1.07 | 1.02 | 0.88 | 0.37 | Malignant neoplasm of breast (174) |
| 532 | 101 | 126 | 104 | 139 | 108 | 1.03 | 1.07 | 0.63 | 0.21 |  |
| 66 | 101 | 11 | 92 | 19 | 103 | 0.91 | 1.02 | 0.40 | 0.45 | Malignant neoplasm of corpus uteri (182) |
| 29 | 97 | 7 | 131 | 5 | 58 | 1.35 | 0.60 | 0.79 | 0.88 |  |
| 301 | 101 | 70 | 102 | 78 | 104 | 1.01 | 1.03 | 0.54 | 0.39 | Malignant neoplasm of ovary (183) |
| 123 | 91 | 30 | 98 | 32 | 92 | 1.08 | 1.01 | 0.67 | 0.46 |  |
| 7 | 71 | 5 | 0 | 0 | 0 |  |  |  |  | Malignant neoplasm of testis (186) |
| 6 | 131 | 3 | 0 | 0 | 0 |  |  |  |  |  |
| 266 | 106 | 48 | 98 | 81 | 112 | 0.92 | 1.06 | 0.30 | 0.31 | Malignant neoplasm of kidney and other and unspecified urinary organs (189) |
| 146 | 104 | 28 | 102 | 46 | 119 | 0.98 | 1.14 | 0.47 | 0.18 |  |
| 106 | 95 | 18 | 83 | 42 | 127 | 0.87 | 1.34 | 0.29 | 0.03 |  |
| 74 | 116 | 10 | 80 | 25 | 140 | 0.69 | 1.21 | 0.12 | 0.17 |  |
| 17 | 83 | 4 | 69 | 4 | 76 | 0.83 | 0.92 | 0.38 | 0.54 | Malignant neoplasm of the thyroid gland (193) |
| 35 | 86 | 12 | 106 | 5 | 61 | 1.23 | 0.71 | 0.77 | 0.77 |  |
| 12 | 131 | 2 | 77 | 2 | 87 | 0.59 | 0.66 | 0.24 | 0.69 |  |
| 18 | 97 | 5 | 98 | 3 | 103 | 1.01 | 1.06 | 0.54 | 0.43 |  |
| 656 | 101 | 145 | 106 | 179 | 102 | 1.05 | 1.01 | 0.72 | 0.44 | Malignant neoplasms of lymphoid, haematopoietic and related tissue (200-208) |
| 545 | 91 | 106 | 91 | 139 | 86 | 1.00 | 0.95 | 0.51 | 0.74 |  |
| 289 | 99 | 61 | 100 | 75 | 94 | 1.01 | 0.95 | 0.54 | 0.67 |  |
| 250 | 91 | 42 | 80 | 67 | 90 | 0.88 | 0.99 | 0.20 | 0.53 |  |
| 594 | 97 | 140 | 96 | 196 | 112 | 0.99 | 1.15 | 0.46 | 0.02 | Diabetes mellitus (250) |
| 1049 | 107 | 298 | 101 | 271 | 126 | 0.94 | 1.18 | 0.16 | 0.00 |  |
| 279 | 102 | 65 | 99 | 103 | 130 | 0.97 | 1.27 | 0.41 | 0.01 |  |
| 469 | 104 | 145 | 108 | 117 | 117 | 1.04 | 1.13 | 0.68 | 0.10 |  |
| 11156 | 111 | 3240 | 105 | 2266 | 116 | 0.95 | 1.05 | 0.00 | 0.02 | Diseases of the circulatory system (390-459) |
| 13757 | 111 | 3583 | 105 | 3166 | 115 | 0.95 | 1.04 | 0.00 | 0.02 |  |
| 5253 | 117 | 1502 | 108 | 1083 | 122 | 0.92 | 1.04 | 0.00 | 0.08 |  |
| 6705 | 117 | 1672 | 107 | 1606 | 127 | 0.91 | 1.09 | 0.00 | 0.00 |  |
| 7632 | 108 | 2006 | 98 | 1662 | 113 | 0.91 | 1.05 | 0.00 | 0.03 | Heart diseases (390-429) |
| 8976 | 107 | 2062 | 98 | 2278 | 115 | 0.92 | 1.07 | 0.00 | 0.00 |  |
| 3534 | 112 | 926 | 101 | 783 | 117 | 0.90 | 1.04 | 0.00 | 0.11 |  |
| 4304 | 111 | 965 | 100 | 1119 | 122 | 0.90 | 1.10 | 0.00 | 0.00 |  |
| 4290 | 111 | 1150 | 100 | 876 | 118 | 0.90 | 1.06 | 0.00 | 0.04 | Ischaemic heart diseases (410-414) |
| 3914 | 111 | 871 | 97 | 892 | 119 | 0.87 | 1.07 | 0.00 | 0.02 |  |
| 1933 | 113 | 506 | 98 | 412 | 122 | 0.87 | 1.08 | 0.00 | 0.06 |  |
| 1876 | 115 | 390 | 96 | 446 | 128 | 0.83 | 1.11 | 0.00 | 0.01 |  |
| 1534 | 114 | 549 | 110 | 285 | 124 | 0.96 | 1.09 | 0.20 | 0.08 | Diseases of the digestive system (520-579) |
| 1327 | 113 | 367 | 115 | 327 | 127 | 1.02 | 1.12 | 0.63 | 0.02 |  |
| 766 | 127 | 275 | 123 | 140 | 134 | 0.97 | 1.06 | 0.30 | 0.26 |  |
| 645 | 120 | 200 | 138 | 147 | 123 | 1.15 | 1.02 | 0.97 | 0.38 |  |

## Notes

### Competing Interest Statement

The authors have declared no competing interest.

### Author Declarations

The study was approved by the Ethics Committee at the Romagna Cancer Institute (ID: IRST100.37).

### Summary of Updates

Supplementary Table S4 added in .xlsx format.

